# Whole genome analysis of plasma fibrinogen reveals population-differentiated genetic regulators with putative liver roles

**DOI:** 10.1101/2023.06.07.23291095

**Authors:** Jennifer E Huffman, Jayna Nicolas, Julie Hahn, Adam S Heath, Laura M Raffield, Lisa R Yanek, Jennifer A Brody, Florian Thibord, Laura Almasy, Traci M Bartz, Lawrence F. Bielak, Russell P Bowler, Germán D Carrasquilla, Daniel I Chasman, Ming-Huei Chen, David B Emmert, Mohsen Ghanbari, Jeffery Haessle, Jouke-Jan Hottenga, Marcus E Kleber, Ngoc-Quynh Le, Jiwon Lee, Joshua P Lewis, Ruifang Li-Gao, Jian’an Luan, Anni Malmberg, Massimo Mangino, Riccardo E Marioni, Angel Martinez-Perez, Nathan Pankratz, Ozren Polasek, Anne Richmond, Benjamin AT Rodriguez, Jerome I Rotter, Maristella Steri, Pierre Suchon, Stella Trompet, Stefan Weiss, Marjan Zare, Paul Auer, Michael H Cho, Paraskevi Christofidou, Gail Davies, Eco de Geus, Jean-François Deleuze, Graciela E Delgado, Lynette Ekunwe, Nauder Faraday, Martin Gögele, Andreas Greinacher, Gao He, Tom Howard, Peter K Joshi, Tuomas O Kilpeläinen, Jari Lahti, Allan Linneberg, Silvia Naitza, Raymond Noordam, Ferran Paüls-Vergés, Stephen S Rich, Frits R Rosendaal, Igor Rudan, Kathleen A Ryan, Juan Carlos Souto, Frank JA van Rooij, Heming Wang, Wei Zhao, Lewis C Becker, Andrew Beswick, Michael R Brown, Brian E Cade, Harry Campbell, Kelly Cho, James D Crapo, Joanne E Curran, Moniek PM de Maat, Margaret Doyle, Paul Elliott, James S Floyd, Christian Fuchsberger, Niels Grarup, Xiuqing Guo, Sarah E Harris, Lifang Hou, Ivana Kolcic, Charles Kooperberg, Cristina Menni, Matthias Nauck, Jeffrey R O’Connell, Valeria Orrù, Bruce M Psaty, Katri Räikkönen, Jennifer A Smith, Jose Manuel Soria, David J Stott, Astrid van Hylckama Vlieg, Hugh Watkins, Gonneke Willemsen, Peter Wilson, Yoav Ben-Shlomo, John Blangero, Dorret Boomsma, Simon R Cox, Abbas Dehghan, Johan G Eriksson, Edoardo Fiorillo, Myriam Fornage, Torben Hansen, Caroline Hayward, M. Arfan Ikram, J Wouter Jukema, Sharon LR Kardia, Leslie A Lange, Winfried März, Rasika A Mathias, Braxton D Mitchell, Dennis O Mook-Kanamori, Pierre-Emmanuel Morange, Oluf Pedersen, Peter P Pramstaller, Susan Redline, Alexander Reiner, Paul M Ridker, Edwin K Silverman, Tim D Spector, Uwe Völker, Nick Wareham, James F Wilson, Jie Yao, VA Million Veteran Program, NHLBI Trans-Omics for Precision Medicine (TOPMed) Consortium, David-Alexandre Trégouët, Andrew D Johnson, Alisa S Wolberg, Paul S de Vries, Maria Sabater-Lleal, Alanna C Morrison, Nicholas L Smith

**Affiliations:** Palo Alto VA Institute for Research, VA Palo Alto Heath Care System, Palo Alto, CA, USA; MAVERIC, VA Boston Healthcare System, Boston, MA, USA; Department of Genetics, University of North Carolina at Chapel Hill, Chapel Hill, NC, USA; Human Genetics Center, Department of Epidemiology, Human Genetics, and Environmental Sciences, School of Public Health, The University of Texas Health Science Center at Houston, Houston, TX, USA; Department of Medicine, Johns Hopkins University School of Medicine, Baltimore, MD, USA; Cardiovascular Health Research Unit, Department of Medicine, University of Washington, Seattle, WA, USA; National Heart Lung and Blood Institute, Division of Intramural Research, Population Sciences Branch, The Framingham Heart Study, Framingham, MA, USA; Department of Biomedical and Health Informatics, Children’s Hospital of Philadelphia, Philadelphia, PA, USA; Department of Biostatistics, University of Washington, Seattle, WA, USA; Department of Epidemiology, School of Public Health, University of Michigan, Ann Arbor, MI, USA; National Jewish Health, Denver, CO, USA; Novo Nordisk Foundation Center for Basic Metabolic Research, Faculty of Health and Medical Sciences, University of Copenhagen, Copenhagen, Denmark; Institute of Environmental Medicine, Karolinska Institutet, Stockholm, Sweden; Division of Preventive Medicine, Brigham and Women’s Hospital, Harvard Medical School, Boston, MA, USA; Institute for Biomedicine (affiliated to the University of Lübeck), Eurac Research, Bolzano, Italy; Department of Epidemiology, Erasmus MC University Medical Center, Rotterdam, the Netherlands; Public Health Sciences Division, Fred Hutchinson Cancer Center, Seattle, WA, USA; Netherlands Twin Register, Department of Biological Psychology, Vrije Universiteit, Amsterdam, the Netherlands; SYNLAB MVZ für Humangenetik Mannheim, Mannheim, Germany; Vth Department of Medicine, Medical Faculty Mannheim, Heidelberg University, Mannheim, Germany; Unit of Genomics of Complex Disease, Institut d’Investigació Biomèdica Sant Pau (IIB SANT PAU), Barcelona, Spain; Department of Medicine, Brigham and Women’s Hospital, Harvard Medical School, Boston, MA, USA; Department of Medicine, University of Maryland, Baltimore, MD, USA; Department of Clinical Epidemiology, Leiden University Medical Center, Leiden, the Netherlands; MRC Epidemiology Unit, University of Cambridge School of Clinical Medicine, Cambridge, UK; Department of Psychology and Logopedics, University of Helsinki, Helsinki, Finland; Department of Twin Research and Genetic Epidemiology, Kings College London, London, UK; NIHR Biomedical Research Centre, Guy’s and St Thomas’ Foundation Trust, London, UK; Centre for Genomic and Experimental Medicine, Institute of Genetics and Cancer, University of Edinburgh, Edinburgh, Scotland, UK; Department of Laboratory Medicine and Pathology, University of Minnesota Medical School, Minneapolis, MN, USA; Faculty of Medicine, University of Split, Split, Croatia; MRC Human Genetics Unit, Institute of Genetics and Cancer, University of Edinburgh, Edinburgh, Scotland, UK; The Institute for Translational Genomics and Population Sciences, Department of Pediatrics, The Lundquist Institute for Biomedical Innovation at Harbor-UCLA Medical Center, Torrance, CA, USA; Istituto di Ricerca Genetica e Biomedica, CNR, Monserrato-Cagliari, Italy; C2VN, INSERM, INRA, Aix Marseille Univ, Marseille, France; Section of Gerontology and Geriatrics, Department of Internal Medicin, Leiden University Medical Center, Leiden, the Netherlands; Interfaculty Institute for Genetics and Functional Genomics, Department of Functional Genomics, German Center for Cardiovascular Research (DZHK), Partner Site Greifswald, Greifswald, Germany, University Medicine Greifswald, Greifswald, Germany; Maternal-Fetal Medicine Research Center, Shiraz University of Medical Sciences, Shiraz, Iran; Department of Epidemiology and Biostatistics, School of Public Health, Imperial College London, London, UK; Division of Biostatistics, Institute for Health and Equity, and Cancer Center,, Medical College of Wisconsin, Milwaukee, WI, USA; Channing Division of Network Medicine and Division of Pulmonary and Critical Care Medicine, Brigham and Women’s Hospital, Harvard Medical School, Boston, MA, USA; Lothian Birth Cohorts, Department of Psychology, University of Edinburgh, Edinburgh, Scotland, UK; Centre National de Recherche en Génomique Humaine, CEA, Université Paris-Saclay, Evry, France; Jackson Heart Study, University of Mississippi Medical Center, Jackson, MS, USA; Department of Anesthesiology and Critical Care Medicine, Johns Hopkins University School of Medicine, Baltimore, MD, USA; Department of Transfusion Medicine, University Medicine Greifswald, Greifswald, Germany; MRC-PHE Centre for Environment & Health, School of Public Health, Imperial College London, London, UK; Department of Human Genetics, University of Texas Rio Grande Valley School of Medicine, Brownsville, TX, USA; Centre for Global Health Research, Usher Institute for Population Health Sciences and Informatics, University of Edinburgh, Edinburgh, Scotland, UK; Center for Clinical Research and Prevention, Bispebjerg and Frederiksberg Hospital, Copenhagen, Denmark; Department of Public Health Sciences, Center for Public Health Genomics, University of Virginia, Charlottesville, VA, USA; Unit of Thrombosis and Hemostasis, Hospital de la Santa Creu i Sant Pau, Barcelona, Spain; Translational Health Sciences, University of Bristol, Bristol, UK; Department of Medicine, Brigham & Women’s Hospital and Harvard Medical School, Boston, MA, USA; Department of Human Genetics and South Texas Diabetes and Obesity Institute, University of Texas Rio Grande Valley School of Medicine, Brownsville, TX, USA; Department of Hematology, Erasmus MC University Medical Center, Rotterdam, the Netherlands; Department of Pathology & Laboratory Medicine, The University of Vermont Larner College of Medicine, Colchester, VT, USA; UK-DRI Dementia Research Institute, Imperial College London, London, UK; Departments of Medicine and Epidemiology, University of Washington, Seattle, WA, USA; Department of Preventive Medicine, Northwestern University, Chicago, IL, USA; Institute of Clinical Chemistry and Laboratory Medicine, German Center for Cardiovascular Research (DZHK), Partner Site Greifswald, Greifswald, Germany, University Medicine Greifswald, Greifswald, Germany; Department of Epidemiology, University of Washington, Seattle, WA, USA; Department of Health Systems and Population Health, University of Washington, Seattle, WA, USA; Institute for Social Research, Survey Research Center, University of Michigan, MI, USA; Institute of Cardiovascular and Medical Sciences, College of Medical, Veterinary and Life Sciences, University of Glasgow, Glasgow, Scotland, UK; Radcliffe Department of Medicine, University of Oxford, Oxford, UK; VA Atlanta Healthcare System, Decatur, GA, USA; Division of Cardiology, Emory University School of Medicine, Atlanta, GA, USA; Department of Epidemiology, Rollins School of Public Health, Emory University, Atlanta, GA, USA; Poulation Health Sciences, University of Bristol, Bristol, UK; Department of General Practice and Primary Health Care, University of Helsinki, Helsinki, Finland; Folkhälsan Research Centre, Helsinki, Finland; Department of Obstetrics and Gynecology, Yong Loo Lin School of Medicine, National University Singapore, Singapore, Singapore; Brown Foundation Institute of Molecular Medicine, McGovern Medical School, University of Texas Health Science Center at Houston, Houston, TX, USA; Department of Cardiology, Leiden University Medical Center, Leiden, the Netherlands; Netherlands Heart Institute, Utrecht, the Netherlands; Division of Biomedical Informatics and Personalized Medicine, Department of Medicine, University of Colorado Anschutz Medical Campus, Aurora, CO, USA; Synlab Academy, Synlab Holding Deutschland GmbH, Mannheim, Germany; Geriatric Research and Education Clinical Center, Baltimore Veterans Administration Medical Center, Baltimore, MD, USA; Department of Public Health and Primary Care, Leiden University Medical Center, Leiden, the Netherlands; Laboratory of Haematology, La Timone Hospital, Marseille, France; Department of Medicine, Beth Israel Deaconness Medical Center, Boston, MA, USA; University of Bordeaux, Bordeaux Population Health Research Center, INSERM UMR 1219, Bordeaux, France; Department of Pathology and Laboratory Medicine and UNC Blood Research Center, University of North Carolina at Chapel Hill, Chapel Hill, NC, USA; Cardiovascular Medicine Unit, Department of Medicine, Karolinska Institutet, Center for Molecular Medicine, Stockholm, Sweden; Kaiser Permanente Washington Health Research Institute, Kaiser Permanente Washington, Seattle, WA, USA; Seattle Epidemiologic Research and Information Center, Department of Veterans Affairs Office of Research and Development, Seattle, WA, USA

## Abstract

Genetic studies have identified numerous regions associated with plasma fibrinogen levels in Europeans, yet missing heritability and limited inclusion of non-Europeans necessitates further studies with improved power and sensitivity. Compared with array-based genotyping, whole genome sequencing (WGS) data provides better coverage of the genome and better representation of non-European variants. To better understand the genetic landscape regulating plasma fibrinogen levels, we meta-analyzed WGS data from the NHLBI’s Trans-Omics for Precision Medicine (TOPMed) program (n=32,572), with array-based genotype data from the Cohorts for Heart and Aging Research in Genomic Epidemiology (CHARGE) Consortium (n=131,340) imputed to the TOPMed or Haplotype Reference Consortium panel. We identified 18 loci that have not been identified in prior genetic studies of fibrinogen. Of these, four are driven by common variants of small effect with reported MAF at least 10% higher in African populations. Three (*SERPINA1, ZFP36L2*, and *TLR10)* signals contain predicted deleterious missense variants. Two loci, *SOCS3* and *HPN*, each harbor two conditionally distinct, non-coding variants. The gene region encoding the protein chain subunits (*FGG;FGB;FGA*), contains 7 distinct signals, including one novel signal driven by rs28577061, a variant common (MAF=0.180) in African reference panels but extremely rare (MAF=0.008) in Europeans. Through phenome-wide association studies in the VA Million Veteran Program, we found associations between fibrinogen polygenic risk scores and thrombotic and inflammatory disease phenotypes, including an association with gout. Our findings demonstrate the utility of WGS to augment genetic discovery in diverse populations and offer new insights for putative mechanisms of fibrinogen regulation.

**Key Points:**

- Largest and most diverse genetic study of plasma fibrinogen identifies 54 regions (18 novel), housing 69 conditionally distinct variants (20 novel).
- Sufficient power achieved to identify signal driven by African population variant.
- Links to (1) liver enzyme, blood cell and lipid genetic signals, (2) liver regulatory elements, and (3) thrombotic and inflammatory disease.

## Introduction

Fibrinogen is a critical coagulation factor and acute phase reactive protein. Under normal conditions, fibrinogen is abundant in circulation, yet during the acute phase inflammatory response, interleukin-(IL)-6 and IL-1 mediate transcriptional cascades which increase circulating fibrinogen levels up to 3-fold above baseline^1,2^. Fibrinogen measures are a clinical predictor of thrombotic diseases, including coronary heart disease, myocardial infarction, venous thromboembolism, and ischemic stroke^3,4^. While animal models have shown a causative relationship between fibrinogen and thrombosis^5,6^, this has been difficult to confirm using Mendelian Randomization^7–9^.

Circulating fibrinogen levels are estimated to be 30-50% heritable^10,11^, and heterogeneous across diverse populations^12–14^. Individuals identifying as African American have higher reported baseline levels of fibrinogen^12–14^, with one study suggesting higher fibrinogen heritability in African ancestral populations (44%) compared with European and other populations (28%)^15^. While genome-wide and exome-wide sequencing studies have identified several loci associated with fibrinogen measures, these variants explain a maximum of 3.7% of variance in European populations^7,16,17^. Little is known regarding genetic regulation of fibrinogen across diverse populations.

Unlike genotyping arrays, which often have better coverage of variants common in European populations, whole genome sequencing (WGS) allows non-targeted genomic interrogation across all populations^18^. Deeper coverage provided by WGS increases confidence in minor allele calls, improving power to detect associations with rare and low frequency variants, and to distinguish multiple signals in the same region^19,20^. Furthermore, deep large-scale WGS-based reference panels improve imputation quality, increasing power derived from genotyped samples in meta-analysis^19,20^. Incorporating more sensitive genomic approaches, such as whole genome sequencing (WGS), in genetic analyses of fibrinogen may reveal additional genetic associations across a range of allele frequencies and effect sizes in diverse populations.

In this study, we performed genome-wide analyses integrating WGS data from NHLBI’s Trans-Omics for Precision Medicine (TOPMed) program^21^ and TOPMed-imputed genotyping data from the Cohorts for Heart and Aging Research in Genomic Epidemiology (CHARGE) Consortium^22^ to identify new genetic variants associated with circulating fibrinogen. To determine putative regulatory mechanisms and prioritize potentially causal genes, we performed *in silico* annotation, colocalization analyses, a transcriptome-wide association study (TWAS), and TWAS fine-mapping. Additionally, we tested the hypothesis that polygenic risk scores (PRS) for fibrinogen are also associated with coagulation and inflammation-related phenotypes in European-ancestry and African-American participants from the VA Million Veteran Program (MVP)^23^.

## Results

### Baseline Characteristics

WGS and fibrinogen measures were available for 32,572 individuals across 13 TOPMed studies. Imputed genetic data and fibrinogen measures were available for 131,340 individuals and 39 additional studies (individuals without WGS from TOPMed studies, and CHARGE studies imputed to either the TOPMed or HRC reference panels). A total of 163,912 individuals were included in the multi-population meta-analysis (**Table 1**). The mean fibrinogen value within TOPMed studies was 3.25 g/L (SD=0.96). Mean values were slightly higher in African ancestry individuals compared to European ancestry individuals, but the confidence intervals overlapped [AFR=3.84 (0.98); EUR=3.26 (0.91)].

### Single Variant Analysis & Aggregate Tests

Multi-population, single-variant meta-analysis identified 54 genetic loci associated with fibrinogen levels (**Figure 2, Supplemental Table 1**). Among these, 18 loci have not been identified in prior genetic studies of fibrinogen (**Table 2**). The *TM6SF2* locus has been identified in a multi-phenotype analysis leveraging fibrinogen and coronary artery disease variants^24^, but not in a GWAS of fibrinogen alone. Approximate conditional analysis in GCTA revealed 6 loci harboring conditionally distinct lead variants **(Table 3; Supplemental Table 2**). At 4 loci with previously reported fibrinogen associations - *FGG, PDLIM4, RPL22L1*, and *RAB37* - we detected 7, 5, 3, and 2 distinct lead variants, respectively, exceeding the maximum at any prior report. The LD between conditionally distinct lead variants and previously published top SNPs is presented in **Supplemental Table 3**. Among newly associated loci, *SOCS3* and *HPN* were found to harbor distinct lead variants.

**Figure 1.**
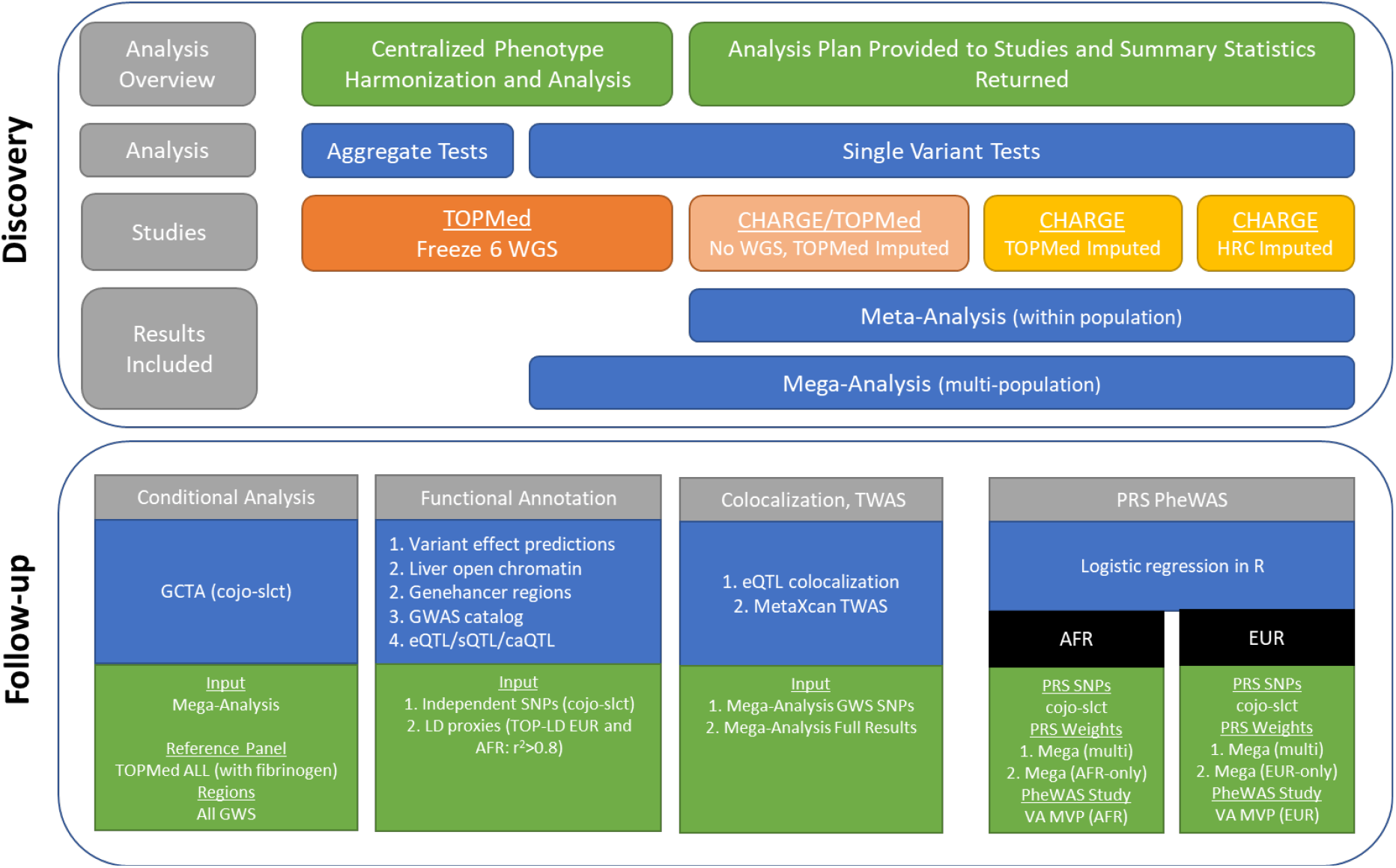
Study Design.

**Figure 2.**
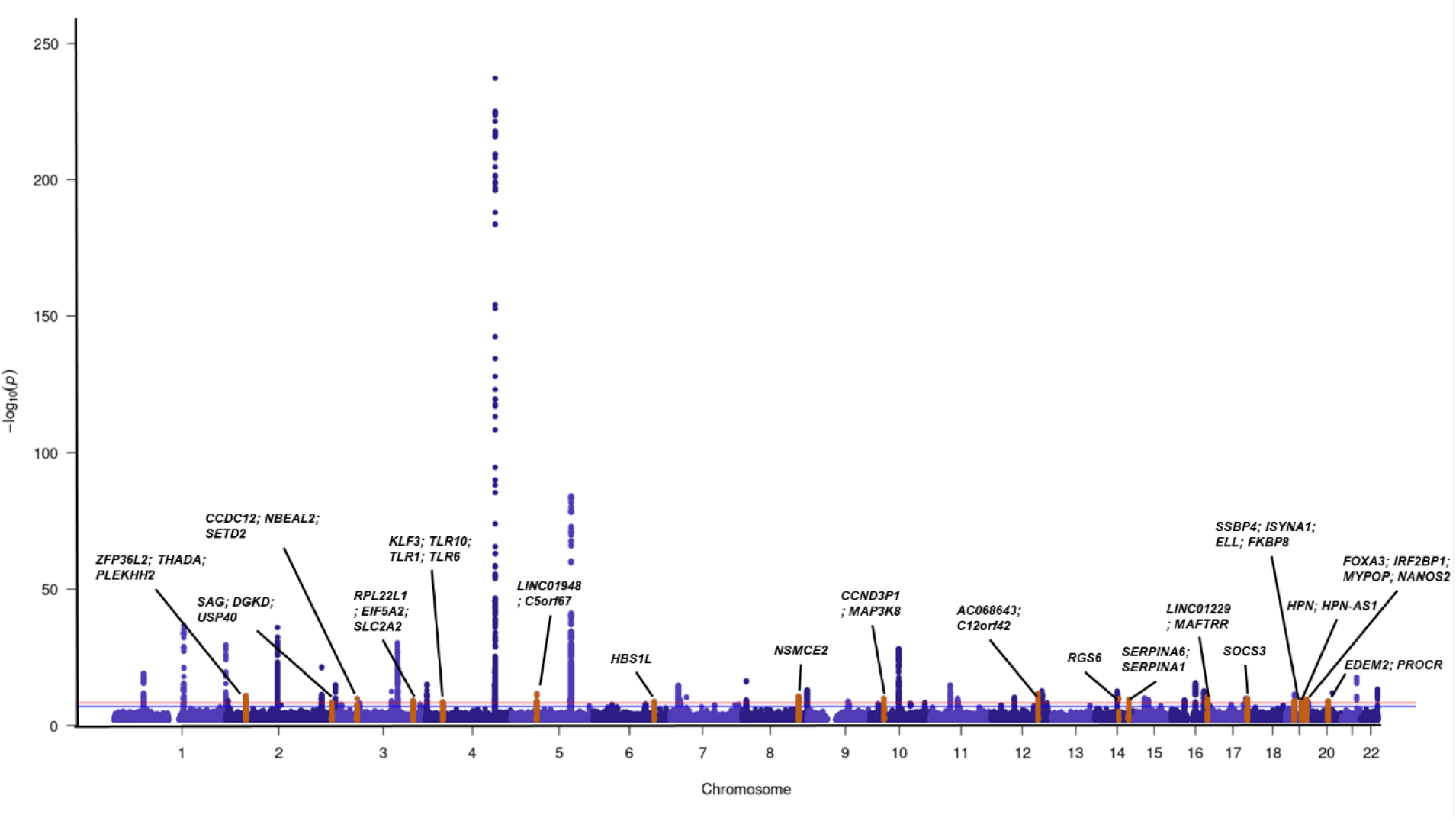
Mega-Analysis Single Variant Results.
Orange peaks are novel associated regions and are labelled with the names of genes in the region. Red line indicates genome-wide significance (p<5E-09) and blue line indicates suggestive association (p<1E-07).

**Figure 3.**
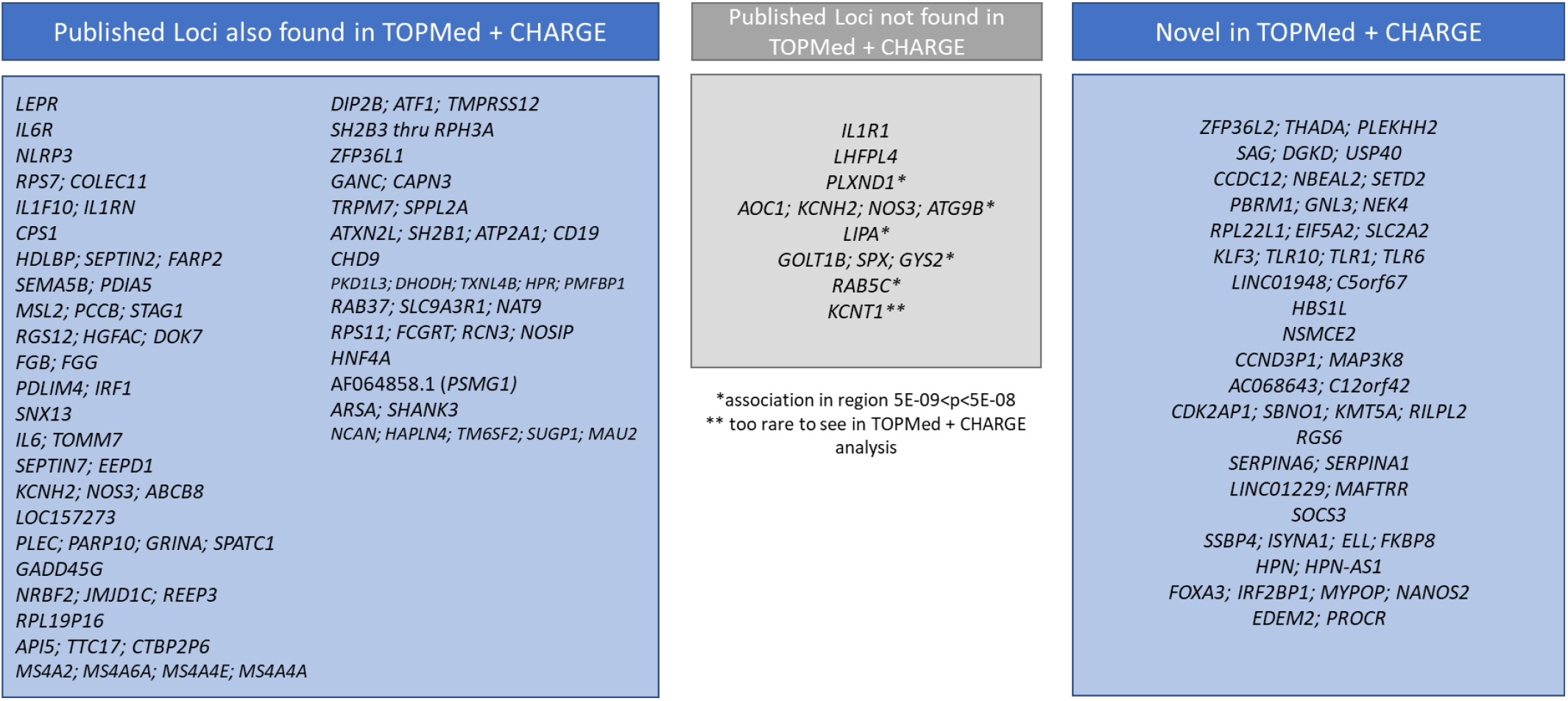
Comparison of Mega-Analysis Single Variant Results to Previous Publications.

Among the 69 conditionally distinct lead variants associated with fibrinogen, 5 variants (rs10936662, rs28577061, rs11077357, rs7507218, rs1672981) have a minor allele at least 10% more frequent in African ancestral populations compared to European (**Supplemental Table 2**). At the *FGG* locus, GCTA identified a conditionally distinct lead variant (rs28577061), which has not been associated with fibrinogen in prior studies and is common in African (MAF= 0.1801) but extremely rare (MAF= 0.008 in EUR) or absent in other ancestral populations in TOPMed. At 4 loci - *RPL22L1, HPN, SOCS3*, and *SSPB4 -* the primary lead variants are common in all assessed populations, but increased frequency in African ancestry populations. Together, the 69 independent variants discovered explain 4.8% of the phenotypic variance for circulating fibrinogen across populations.

Aggregate tests using low-frequency and rare TOPMed WGS variants yielded associations in the fibrinogen gene cluster region – *FGG* was significant when aggregating (1) loss-of-function (LOF) variants, and (2) LOF and deleterious missense variants whereas *FGA* was only significant when aggregating LOF variants and *FGB* with LOF, deletions, and missense variants. No genes were significant when aggregating all low-frequency and rare variants in the coding, promoter, and enhancer regions (**Supplemental Table 4**).

### Variant annotation

To characterize each genetic signal associated with fibrinogen, we queried all conditionally distinct variants and their LD proxies (r^2^>0.8 in TOPMed-based European and/or African ancestry reference panels)^21,25^, in multiple publicly available datasets. The majority of signals (defined as the conditionally distinct variant and LD-proxies) we identified contain previously reported lead variants for liver-enzyme measures, lipid measures, and/or blood-cell traits. Notably, 23 signals contain a previously reported lead variant for C-Reactive Protein (CRP).

Variant effect prediction (VEP)^26^ shows that 18 signals contain at least one missense variant. Additionally, 13 signals contain variants with CADD PHRED scores exceeding 20, indicating variant status in the top 1% of predicted deleterious mutations. *SERPINA1, ZFP36L2*, and *TLR10*, stand out as newly associated loci in this category. We also observed that several signals overlap potential regulatory regions in liver. 52 signals (45 loci) contain at least 1 variant mapping to a “consensus” region of open chromatin in liver tissue samples. 42 signals overlap a GeneHancer regulatory element reported active in HepG2 hepatocytes. 5 signals harbor previously reported liver chromatin accessibility quantitative trait loci (QTL) variants, 6 signals hold GTEx liver expression QTL, and the primary signal at the *HPN* locus contains a GTEx liver splice QTL variant. A summary of these annotations for each signal can be found in **Supplemental Table 5**. Full results of this analysis are in **Supplemental Table 6**.

### Colocalization

Variant-level colocalization analysis in fastENLOC identified 153 variant-tissue pairs with evidence (SCP>0.1) for a shared genetic basis with GTEx expression QTLs for 93 distinct genes in aortic artery, tibial artery, coronary artery, liver, and/or whole blood (**Supplemental Table 7**). Regional analysis in the same set of tissues found 46 region-tissue pairs, implicated in regulation of 41 distinct genes, with evidence for statistical colocalization with fibrinogen regions (RCP>0.5) (**Supplemental Table 8**). We note that 5 of the 6 fibrinogen signals harboring sentinel GTEx liver eQTL variants statistically colocalize with the expression QTL signal in fastENLOC, while the *PLEC* eQTL signal was at the significance threshold (RCP = 0.5).

### Transcriptome-wide Association Study

Gene-tissue pairs across 5 tissues (aortic artery, coronary artery, tibial artery, whole blood, and liver) were included in MetaXcan TWAS analyses for fibrinogen. Genetically determined expressions of 64 gene-tissue pairs were significantly associated with fibrinogen levels after Bonferroni correction and filtering for models with at least 2 contributing SNPs (**Supplemental Table 9**). 15 gene-tissue pairs identified by TWAS were further prioritized via fine-mapping in FOCUS (**Supplemental Table 10**). *TNKS* was the only gene prioritized by both TWAS and fine-mapping in liver-analyses. Notably, this gene was further implicated in fibrinogen regulation through variant and region-based colocalization analysis using GTEx liver expression data (**Supplemental Table 8**). In whole blood analyses, TWAS and fine-mapping prioritized *AFT1, C5orf56, FGB, MS4A4E, SLC22A5*. Of these, *AFT1* was further supported by variant-level colocalization analysis (**Supplemental Table 7**).

### Phenomewide Association Study (PheWAS)

Three fibrinogen PRSs were tested for association with 1,426 ICD-based PheCodes in EUR and 1,061 PheCodes in AFR participants of the VA MVP **(Supplemental Table 11)**. Each score reached Bonferroni significance with multiple thrombotic and inflammatory disease phenotypes (n_EUR_=13 PheCodes, n_AFR_=1 PheCode; **Table 4**). Top associations included venous thromboembolism and gout.

To further assess which variants within the PRS may drive individual associations, we queried each variant against the significant PheCodes (**Supplemental Table 12**). As anticipated, for “Circulatory System” and “Hematopoietic” PheCodes, most of the signal came from variants in the fibrinogen gene cluster, whereas significant variants across the PRS were associated with gout and dermatologic traits. Sensitivity analyses with PRS removing variants either in the *FGG* gene region (associated with gamma prime fibrinogen) or in the full fibrinogen gene cluster did not substantially change the odds ratio, but did impact the significance (**Supplemental Table 13**). PheCodes relating to gout (phe_274, phe_274_1) and superficial cellulitis and abscess (phe_1089) became more significant, whereas PheCodes relating to coagulation defects and hypercoagulable state (phe_286, phe_286_8, phe_286_81) became less significant.

## Discussion

Here, we used WGS and genotype data from diverse participants to identify 69 conditionally distinct genetic variants from 54 loci associated with circulating fibrinogen across populations. Our results corroborate previous reports that fibrinogen is highly polygenic and advance the field by identifying new variant associations, including rare variants and variants most prevalent in underrepresented populations. Based on results of *in silico* characterization, we suggest some of the new genetic regulators we identified may act directly to regulate coagulation factors (such as factor VII, factor XI), while some may impact fibrinogen broadly through pathways altering liver metabolism, inflammation, and immune function, as reflected in the broad overlap between fibrinogen associated signals and prior analyses for liver-related traits (such as lipids and CRP) and blood-cell and immune-cell counts.

### Population-differentiated variants among new associations

We identified several novel genetic associations driven by variants with higher allele frequencies in non-European populations, validating that our approach improved detection of putative genetic regulators which may be most common in underrepresented populations. To our knowledge, we identified the first common variant-fibrinogen association driven almost entirely by African ancestry participants at an intergenic region near *FGG*. Previous African ancestry driven variants were rare in African and not present in European populations^17^. Additionally, expanded representation of both African and European populations allowed us to detect 4 new signals of small effect, led by common variants substantially (>10%) more frequent in those with African ancestry. Detection of population-differentiated associations indicates that for variants with higher predicted effects, such as those within the fibrinogen gene cluster, expanded sample sizes are now reaching power to detect variant associations driven by underrepresented populations, although improved representation is still needed. We further identified 4 common European-driven signals that are rare or uncommon in African ancestry populations, suggesting that genomic signals for circulating fibrinogen are not saturated in any ancestral population^27,28^, as reflected by the relatively modest variance explained (4.8%) versus estimates of total fibrinogen heritability (28-44%)^10,11,15^. With improved representation of all ancestries, genomic studies will likely identify additional common and rare genetic signals across all populations.

### Newly identified potential coagulation pathway interactors

Three novel regions (*SERPINA1, ZFP36L2*, and *TLR10*) harbor deleterious missense variants in genes with plausible connections to coagulation and/or inflammation. In the *SERPINA1* gene, we identified a rare missense variant (rs17580) with a high CADD PHRED score (32). *SERPINA1* encodes alpha-1-antitrypsin, which protects other proteins and tissues from serine protease degradation via direct binding inhibition^29^. Like fibrinogen, alpha-1-antitrypsin is synthesized in hepatocytes in response to IL-6 and IL-1 inflammatory pathways^30^. Severe alpha-1-antitrypsin deficiency is associated with liver dysfunction and fibrotic lung disease related to a lack of inhibition of neutrophil elastase^31^. The association of *SERPINA1* with fibrinogen levels may reflect SERPINA1 involvement in inflammatory pathways, or a more direct relationship between these proteins. While *in vitro* assays have suggested alpha-1-antitrypsin can bind to plasma fibrinogen and be incorporated into fibrin networks^29^, little is known about the functional implications of these interactions and whether it also influences fibrin(ogen) degradation.

ZFP36L2 and TLR10 are broadly linked to inflammatory pathways. Although ZFP36L2 is largely uncharacterized, the *ZFP36L2* missense variant we observed has previously been associated with blood cell traits^32–35^, and studies in mice show ZFP36L2 regulates blood cell development and adipogenesis^36^. Proteins of the ZFP36 family regulate transcript abundance through binding AU-rich elements to target transcripts for decay^37^. Notably, ZFP36 (tristetraprolin) family proteins are known to modulate transcript abundance of the inflammatory cytokine TNF-a^38^, which induces both the coagulation cascade and complement system^39,40^. *TLR10* encodes a toll-like receptor (TLR) protein. While TLR activation generally contributes to inflammation, immune responses, and activation of coagulation cascades, little is known about TLR10 specifically^41^. Although TLR10 may exert a protective effect against inflammation^42,43^, more work is needed to elucidate mechanistic links between this protein and coagulation.

### Putative fibrinogen regulatory signals

While the missense variants highlight intriguing new gene targets for future functional studies, most signals we identified are driven by common non-coding variants with small effect sizes. Many of these non-coding signals overlap HepG2-active regulatory regions, and a subset of signals overlap liver chromatin accessibility, expression, and/or splice QTL signals, suggesting these variants may exert subtle regulatory effects on circulating fibrinogen levels. Notably, at 2 newly associated loci, *HPN* and *SOCS3*, we identified 2 conditionally distinct variants, each with distinct functional annotations.

At the *HPN* locus, the primary signal, which maps to an intronic region of *HPN*, contains a liver splice QTL for *HPN. HPN* encodes hepsin, a type II transmembrane serine protease with “enhanced” expression in the liver (HPA), believed to function in macromolecular metabolism^44^. Interestingly, studies in zebrafish and mammalian cell lines suggest hepsin activates coagulation factor VII^45,46^. Although mouse studies have not conclusively demonstrated hepsin’s involvement in coagulation, these studies have established hepsin’s involvement in regulating liver metabolism via hepatocyte growth factor (HGF) and Met signaling pathways^44,47–49^. The secondary signal at this locus maps to an intergenic region and contains a liver expression QTL for a nearby gene, *TMEM147. TMEM147*, which was also prioritized in liver, whole blood, and aortic artery TWAS analyses, encodes a widely-expressed endoplasmic reticulum transmembrane protein implicated in various metabolic processes, including calcium transport^50^.

Although it is unclear whether the two signals in this region influence fibrinogen through altered splicing and expression patterns of *HPN* and *TMEM147* in the liver, these signals provide a compelling starting point for future functional studies.

Similarly, among the two independent, common-variant signals in the *SOCS3*; *PGS1* locus on chromosome 17, the secondary signal houses a previously identified sentinel variant for altered chromatin accessibility in liver tissue. This variant is of particular interest given its proximity to *SOCS3*, a “Suppressor of Cytokine Signaling” known to act upstream of IL-6 in the acute-phase response pathway that induces fibrinogen^2^. It is possible that the secondary signal at this locus is capturing genetic variation which modulates the accessibility of the *SOCS3* gene, and potentially other nearby genes, in liver or other tissues, leading to downstream impacts on fibrinogen levels. *SOCS3* methylation was recently associated with circulating fibrinogen levels in an epigenomewide association study in the CHARGE consortium^51^ with the associated probe within 11kb of our best *SOCS3* GWAS SNP, reinforcing the idea that chromatin accessibility may be at play.

### PheWAS

Results of the PheWAS in MVP yielded expected associations with venous thromboembolism and several hematopoietic traits. However, the direction of the association within bleeding and thrombosis phenotypes was often opposite to the direction we expected given total fibrinogen’s procoagulant function (i.e., typically increased genetically-predicted fibrinogen is associated with decreased risk of bleeding and increased risk of thrombosis). Although surprising, this finding is in line with a previous Mendelian randomization study, which observed an inverse association between total fibrinogen levels with venous thromboembolism risk^9^. We tested the hypothesis that this inverse association in our PheWAS was driven by the alternatively-spliced gamma prime fibrinogen isoform, which has established anticoagulant properties^52^, but removing the *FGG* locus SNPs from the tested PRS did not alter the direction of effect we observed.

Another unexpected finding from the PheWAS was a positive association between genetically-predicted total fibrinogen and gout. Interestingly, this finding is in line with emerging literature, which has shown that patients with active gout have increased thrombin generation markers^53,54^. Given the extensive overlap between our signals and previously reported GWAS signals for phenotypes reflecting liver health and inflammation - such as liver enzyme, lipid measures, and CRP measure - we suggest that this association may be capturing variant impacts on broader liver metabolic pathways.

### Conclusion

In conclusion, we identified 18 novel loci, collectively harboring 20 distinct variants, associated with fibrinogen measure. We report the first African-variant driven fibrinogen association, and several additional associated variants with population-differentiated genetic architecture. Furthermore, we demonstrate overlap between these signals and liver regulatory elements, as well as GWAS phenotypes reflecting altered liver metabolism and inflammation. Future studies investigating co-regulation and epistatic effects will likely provide new insight on the shared genetic architecture, and biological interplay, of hemostasis and inflammation.

## Methods

### Design and Study Population

To investigate the genetic architecture of circulating fibrinogen, we performed a multi-population genome wide association study, followed by transcriptome-wide (TWAS) and phenome-wide association (PheWAS) studies. An overview of the study design is in **Figure 1**. These analyses were undertaken within the setting of NHLBI’s TOPMed Program and the CHARGE Consortium Hemostasis Working Group. In total, 161,643 participants contributed to primary genomic analyses, including 11,283 African-ancestry (AFR), 741 Asian-ancestry (ASN), 149,619 European-ancestry (EUR), and 2,061 Hispanic (HIS) participants. Assignment to 1 of these 4 groups was determined by each study internally.

Studies often used some combination of self-reported race or ethnicity data (which in many cases was used for stratification at the genotyping stage, making future pooled analysis challenging) and comparison of ancestry PCs to commonly used reference panels such as 1000G (often with exclusion of any extreme outliers). We acknowledge that this is not concordant with the most up to date standards for defining ancestry clusters based on similarity to reference panels^55^ but do not have access to individual level data for most participants and are thus reliant on these study assignments. Details of the 40 participating studies are in **Table 1** and the **Supplemental Data**.

### Phenotyping and Harmonization

Fibrinogen was measured in g/L. Most studies measured clottable fibrinogen using the Clauss method^56^, while the remaining 7 studies used a variety of approaches to measure fibrinogen antigen, including nephlometry and ELISA. Study-specific phenotyping methods can be found in **Table 1** and the **Supplemental Data**. Measures of plasma fibrinogen for TOPMed studies were harmonized to ensure they were in the same units (g/L) and did not have unexpected distributions or an excess of outliers. Data were then uploaded to Analysis Commons^57^ for centralized genetic analysis.

### Whole-genome Sequencing of TOPMed Participants

TOPMed WGS methods were described previously^21^. Briefly, WGS was conducted at six sequencing centers (mean depth >30X, Illumina HiSeq X Ten instruments). Joint variant discovery and genotype calling were conducted by the TOPMed Informatics Research Center (IRC) across all TOPMed studies using the GotCloud pipeline, resulting in a single genotype call set encompassing all of TOPMed (TOPMed Freeze 6). Variant quality control was also performed centrally by the TOPMed IRC. Sample quality control was performed by the TOPMed Data Coordinating Center (DCC). Further details are in the **Supplemental Data**.

### Genotype Imputation of non-TOPMed Studies

Genotype array data for CHARGE studies and for participants without WGS from TOPMed studies were imputed using standard methods to the densest available imputation panel. A total of 35 studies imputed to the TOPMed Freeze 5b reference panel^21^ and 4 to the Haplotype Reference Consortium (HRC) reference panel^58^.

### Genome-wide Association Analyses and Meta-analyses

TOPMed WGS genetic analyses were conducted using inverse normalized and rescaled residuals adjusting for age, sex, population group*study, TOPMed sequencing phase, study-specific parameters, 11 ancestry-informative principal components, and a kinship matrix. Single variant and aggregate gene-based tests were implemented using the SMMAT function of GENESIS on the Analysis Commons cloud computing platform^57,59^. Aggregate tests included only variants with minor allele frequency (MAF) <5% and minor allele count (MAC) ≥1 and used 3 strategies for variant selection: (1) loss of function (LOF), (2) LOF and deleterious missense (LDM), and (3) coding, enhancer, and promoter variants.

Studies without sequencing data undertook single variant analyses within each population group using their software of preference and the same model described for TOPMed genetic analyses. Summary statistics were provided for central meta-analysis. Quality control of the study-specific single variant GWAS summary statistics was undertaken using the EasyQC package^60^ for R. Variants were removed based on the following filtering criteria: estimated minor allele count (minor allele count x imputation quality; eMAC) < 6, absolute effect size (beta) > 5, standard error > 10, sample size < 30, or imputation quality < 0.30. Meta-analysis was completed using GWAMA^61^ with genomic control applied to each study individually but not to the meta-analysis results. Meta-analysis was completed (i) within each population group for just the studies with imputed data, (ii) just the TOPMed WGS studies, and then (iii) combined in a multi-population mega-analysis that included results from analysis of WGS and imputed data. Statistical significance was set at p<5.0E-09^62^.

### Conditional Analysis and Variant Explained

For all genome-wide significant regions from the multi-population mega-analysis, conditional analyses were undertaken using cojo-slct^63^ within GCTA^64^ using all TOPMed WGS samples that contributed to the GWAS as the linkage disequilibrium (LD) reference panel. Percent variance explained was estimated with summary level data reported by GCTA^64^ for the 69 conditionally independent variants from the mega-analysis, using the approximation derived by Shim et al^65^. See **Supplemental Methods** for more details.

### Functional Annotation of Fibrinogen-Associated Variants

#### Variant Effect Prediction

The Ensembl Variant Effect Predictor (VEP) (https://useast.ensembl.org/Tools/VEP)^26^ was used to determine nearest gene and top predicted consequence for each of the 69 conditionally independent variants and their linkage-disequilibrium (LD) proxies (as determined by r^2^>0.8 in TOP-LD European and/or African ancestry reference panels^21,25^). VEP was also used to annotate CADD PHRED^66,67^ and LoFtool^68^-predicted impact scores. For coding variants, SIFT^69^, and PolyPhen^70,71^ scores were obtained through VEP. InterPro was used to determine amino acid substitution^72^.

#### Overlap of Fibrinogen Signals and GWAS Catalog Associations

The NHGRI-EBI GWAS catalog v1.0.2, containing lead, significant (5.0E-08) variants from each uploaded GWAS study was downloaded October 29, 2022 (https://www.ebi.ac.uk/gwas/downloads). The same set of variants used for VEP annotation were queried in the GWAS catalog by rsID. Mapped traits corresponding to related quantitative measures were manually cataloged to broad categories (ie “liver enzymes”, “blood cell traits”, “lipids” etc).

### Colocalization

We performed colocalization to support gene-trait associations using fastENLOC^73,74^. We took the pre-computed GTEx multiple-tissue eQTL annotations and fibrinogen GWAS PIP input to perform fastENLOC for each of the most relevant tissues: liver and whole blood. We considered RCP > 0.5 as strong evidence of colocalization.

### Transcription-wide Association Studies

A transcriptome-wide association study (TWAS) was performed using S-PrediXcan^75^ to identify associations between *cis*-genetic components of gene expression and plasma levels of fibrinogen in mechanistically related tissues, namely artery (aorta, coronary, or tibial), liver, and whole blood^76^. We used the prebuilt prediction models that were based on GTEx v8 multivariate adaptive shrinkage in R to estimate variants’ weight on gene expression levels in chosen tissues^75–77^. Since the reference models were created based on the European-ancestry population, we limited the analysis to GWAS results of European-ancestry individuals only. S-PrediXcan results of the tissues were evaluated using S-MultiXcan^78^. We determined significant TWAS signals using Bonferroni correction for the total number of genes across all models.

### Fine-mapping

To assist in the identification of the causal gene under the GWAS signal, we performed TWAS fine-mapping using FOCUS. FOCUS avoids false TWAS signals caused by co-regulation and pleiotropic effects of variants at GWAS risk loci^79^by modeling marginal TWAS z-scores of all genes in the same region considering variant LD correlations and tagged pleiotropic effects of variants on the trait. Given generated z-scores, posterior inclusion probability (PIP) for a gene to be causal is derived and then used to form a credible set of putative causal genes. In this analysis, we used GTEx v8 MASH-R models as the source of eQTL weights and the European-based PROCARDIS database as the reference for LD correlations. PIP ≥ 0.95 was used as the threshold to determine putative causal genes in FOCUS results.

### MVP PheWAS/Polygenic Risk Scores (PRS) analysis

Polygenic risk scores (PRS) were derived using the independent variants identified by the GCTA analysis described above. Three scores were derived, (i) weighting by the variant beta from the multi-population mega-analysis, (ii) weighting by the variant beta from the European (EUR)-only meta-analysis, and (iii) weighting by the variant beta from the African (AFR) -only meta-analysis. PRS were then standardized to standard deviation units. A phenome-wide association study (PheWAS) was performed for each of the 3 PRS within EUR and AFR participants of the VA Million Veteran Program^23^ (MVP), for ICD-based PheCodes^80^ with at least 500 cases. Logistic regression models were adjusted for age, sex, and the first 5 population-specific principal components and significance determined based on Bonferroni correction for the number of independent PheCodes. EUR had 1,426 PheCodes analyzed with an estimated 965 independent (p_Bonferroni_ = 0.05/965=5.18E-05) and AFR had 1,061 PheCodes with an estimated 690 independent (p_Bonferroni_ = 0.05/690=7.25E-05) (see **Supplemental Data**). Additional sensitivity analyses were completed by creating the PRS removing variants in the *FGG* gene region, or the fibrinogen gene cluster region.

## Supporting information

Main and Supplemental Tables

## Data Availability

Meta-analysis summary statistics will be available through the CHARGE dbGaP accession (phs000930) or upon request to the corresponding author. Individual-level data for TOPMed studies is available through their relevant dbGaP accessions as listed in the TOPMed Omics Support Table in the Supplemental Data.

## Acknowledgements

Molecular data for the Trans-Omics in Precision Medicine (TOPMed) program was supported by the National Heart, Lung and Blood Institute (NHLBI). See the TOPMed Omics Support Table in the **Supplemental Data** for study specific omics support information. Core support including centralized genomic read mapping and genotype calling, along with variant quality metrics and filtering were provided by the TOPMed Informatics Research Center (3R01HL-117626-02S1; contract HHSN268201800002I). Core support including phenotype harmonization, data management, sample-identity QC, and general program coordination were provided by the TOPMed Data Coordinating Center (R01HL-120393; U01HL-120393; contract HHSN268201800001I). We gratefully acknowledge the studies and participants who provided biological samples and data for TOPMed.

CHARGE is supported by R01HL-105756 and the CHARGE and TOPMed Hemostasis Working Groups by R01HL-134894, R01HL-139553, and R01HL-141291.

This research is based on data from the Million Veteran Program, Office of Research and Development, Veterans Health Administration, and was supported by award I01-BX004821.

The study was supported by the Novo Nordisk Foundation (grant number NNF18CC0034900). D-A.T was supported by the EPIDEMIOM-VT Senior Chair of Excellence from the University of Bordeaux initiative of Excellence (IdEX). Additional study-specific funding and acknowledgements can be found in the **Supplemental Data**.

The Genotype-Tissue Expression (GTEx) Project was supported by the Common Fund of the Office of the Director of the National Institutes of Health, and by NCI, NHGRI, NHLBI, NIDA, NIMH, and NINDS.

This publication does not represent the views of the Department of Veteran Affairs, the US National Institutes of Health, the National Heart, Lung and Blood Institute, the US Department of Health and Human Service, or the United States Government.

## Data Availability

Meta-analysis summary statistics are available through the CHARGE dbGaP accession (phs000930) or upon request to the corresponding author. Individual-level data for TOPMed studies is available through their relevant dbGaP accessions as listed in the TOPMed Omics Support Table in the **Supplemental Data**.

## Tables *(located with Supplementary Tables)*

Table 1 – Participating Studies

Table 2 – Novel Regions Associated with Fibrinogen in Single Variant Test

Table 3 – Regions with multiple SNPs selected by GCTA (cojo-slct) using TOPMed multi-population reference panel for LD

Table 4 – Association of TOPMed/CHARGE Fibrinogen Polygenic Risk Scores (PRS) with PheCodes in MVP

