## Supplemental Information, Methods, and Figures for "Whole genome analysis of plasma fibrinogen reveals population-differentiated genetic regulators with putative liver roles"

### Supplemental Data

### Additional Acknowledgements

#### Study-Specific Acknowledgements

**AMISH**

The TOPMed component of the Amish Research Program was supported by NIH grants R01 HL121007, U01 HL072515, and R01 AG18728.

**ARIC**

The Atherosclerosis Risk in Communities study has been funded in whole or in part with Federal funds from the National Heart, Lung, and Blood Institute, National Institutes of Health, Department of Health and Human Services (contract numbers HHSN268201700001I, HHSN268201700002I, HHSN268201700003I, HHSN268201700004I and HHSN268201700005I). The authors thank the staff and participants of the ARIC study for their important contributions.

**CARDIA**

The Coronary Artery Risk Development in Young Adults Study (CARDIA) is conducted and supported by the National Heart, Lung, and Blood Institute (NHLBI) in collaboration with the University of Alabama at Birmingham (HHSN268201800005I & HHSN268201800007I), Northwestern University (HHSN268201800003I), University of Minnesota (HHSN268201800006I), and Kaiser Foundation Research Institute (HHSN268201800004I). CARDIA was also partially supported by the Intramural Research Program of the National Institute on Aging (NIA) and an intra-agency agreement between NIA and NHLBI (AG0005).

**CFS**

The Cleveland Family Study has been supported in part by National Institutes of Health grants [R01-HL046380, KL2-RR024990, R35-HL135818, and R01-HL113338].

**CHS**

This research was supported by contracts HHSN268201200036C, HHSN268200800007C, HHSN268201800001C, N01HC55222, N01HC85079, N01HC85080, N01HC85081, N01HC85082, N01HC85083, N01HC85086, 75N92021D00006, and grants U01HL080295 and U01HL130114 from the National Heart, Lung, and Blood Institute (NHLBI), with additional contribution from the National Institute of Neurological Disorders and Stroke (NINDS). Additional support was provided by R01AG023629 from the National Institute on Aging (NIA). A full list of principal CHS investigators and institutions can be found at CHS-NHLBI.org. The content is solely the responsibility of the authors and does not necessarily represent the official views of the National Institutes of Health.

**COPDGene**

The COPDGene project described was supported by Award Number U01 HL089897 and Award Number U01 HL089856 from the National Heart, Lung, and Blood Institute. The content is solely the responsibility of the authors and does not necessarily represent the official views of the National Heart, Lung, and Blood Institute or the National Institutes of Health. The COPDGene project is also supported by the COPD Foundation through contributions made to an Industry Advisory Board that has included AstraZeneca, Bayer Pharmaceuticals, Boehringer Ingelheim, Genentech, GlaxoSmithKline, Novartis, Pfizer, and Sunovion. A full listing of COPDGene investigators can be found at: http://www.copdgene.org/directory.

**FHS**

The Framingham Heart Study (FHS) acknowledges the support of contracts NO1-HC-25195, HHSN268201500001I and 75N92019D00031 from the National Heart, Lung and Blood Institute and grant supplement R01 HL092577-06S1 for this research. We also acknowledge the dedication of the FHS study participants without whom this research would not be possible. Dr. Vasan is supported in part by the Evans Medical Foundation and the Jay and Louis Coffman Endowment from the Department of Medicine, Boston University School of Medicine.

**GENOA**

Support for GENOA was provided by the National Heart, Lung and Blood Institute (HL054457, HL054464, HL054481, HL119443, and HL087660) of the National Institutes of Health.

**INTER99**

We thank the whole Inter99-staff and all people participating in the study. The Inter99 study was initiated by Torben Jorgensen (principal investigator), Knut Borch-Johnsen (principal investigator on the diabetes part), Troels Thomsen, and Hans Ibsen. The Inter99 study was funded by the Danish Research Councils, Health Foundation, Danish Centre for Evaluation and Health Technology Assessment, Copenhagen County, Danish Heart Foundation, Ministry of Health and Prevention, Association of Danish Pharmacies, Augustinus Foundation, Novo Nordisk, Velux Foundation, Becket Foundation, and Ib Henriksens Foundation.

**JHS**

The Jackson Heart Study (JHS) is supported and conducted in collaboration with Jackson State University (HHSN268201800013I), Tougaloo College (HHSN268201800014I), the Mississippi State Department of Health (HHSN268201800015I) and the University of Mississippi Medical Center (HHSN268201800010I, HHSN268201800011I, and HHSN268201800012I) contracts from the National Heart, Lung, and Blood Institute (NHLBI) and the National Institute on Minority Health and Health Disparities (NIMHD). The authors also wish to thank the staff and participants of the JHS.

**MARTHA**

MARTHA genetic data were funded by the GENMED Laboratory of Excellence (LABEX) on Medical Genomics [ANR-10-LABX-0013], a research program managed by the National Research Agency (ANR) as part of the French Investment for the Future.

**MESA**

MESA and the MESA SHARe projects are conducted and supported by the National Heart, Lung, and Blood Institute (NHLBI) in collaboration with MESA investigators. Support for MESA is provided by contracts 75N92020D00001, HHSN268201500003I, N01-HC-95159, 75N92020D00005, N01-HC-95160, 75N92020D00002, N01-HC-95161, 75N92020D00003, N01-HC-95162, 75N92020D00006, N01-HC-95163, 75N92020D00004, N01-HC-95164, 75N92020D00007, N01-HC-95165, N01-HC-95166, N01-HC-95167, N01-HC-95168, N01-HC-95169, UL1-TR-000040, UL1-TR-001079, and UL1-TR-001420, UL1TR001881, DK063491, and R01HL105756. Funding for SHARe genotyping was provided by NHLBI Contract N02-HL-64278. Genotyping was performed at Affymetrix (Santa Clara, California, USA) and the Broad Institute of Harvard and MIT (Boston, Massachusetts, USA) using the Affymetrix Genome-Wide Human SNP Array 6.0. The authors thank the other investigators, the staff, and the participants of the MESA study for their valuable contributions. A full list of participating MESA investigators and institutes can be found at http://www.mesa-nhlbi.org.

**MVP**

The VA Million Veteran Program (MVP) wishes to acknowledge the following core teams and members:

**MVP Executive Committee**

- Co-Chair: J. Michael Gaziano, M.D., M.P.H.
  VA Boston Healthcare System, 150 S. Huntington Avenue, Boston, MA 02130
- Co-Chair: Sumitra Muralidhar, Ph.D.
  US Department of Veterans Affairs, 810 Vermont Avenue NW, Washington, DC 20420
- Rachel Ramoni, D.M.D., Sc.D., Chief VA Research and Development Officer
  US Department of Veterans Affairs, 810 Vermont Avenue NW, Washington, DC 20420
- Jean Beckham, Ph.D.
  Durham VA Medical Center, 508 Fulton Street, Durham, NC 27705
- Kyong-Mi Chang, M.D.
  Philadelphia VA Medical Center, 3900 Woodland Avenue, Philadelphia, PA 19104
- Philip S. Tsao, Ph.D.
  VA Palo Alto Health Care System, 3801 Miranda Avenue, Palo Alto, CA 94304
- James Breeling, M.D., Ex-Officio
  US Department of Veterans Affairs, 810 Vermont Avenue NW, Washington, DC 20420
- Grant Huang, Ph.D., Ex-Officio
  US Department of Veterans Affairs, 810 Vermont Avenue NW, Washington, DC 20420
- Juan P. Casas, M.D., Ph.D., Ex-Officio
  VA Boston Healthcare System, 150 S. Huntington Avenue, Boston, MA 02130

**MVP Program Office**

- Sumitra Muralidhar, Ph.D.
  US Department of Veterans Affairs, 810 Vermont Avenue NW, Washington, DC 20420
- Jennifer Moser, Ph.D.
  US Department of Veterans Affairs, 810 Vermont Avenue NW, Washington, DC 20420

**MVP Recruitment/Enrollment**

- MVP Cohort Management Director/Recruitment/Enrollment Director, Boston – Stacey B. Whitbourne, Ph.D.; Jessica V. Brewer, M.P.H.
  VA Boston Healthcare System, 150 S. Huntington Avenue, Boston, MA 02130
- VA Central Biorepository, Boston – Mary T. Brophy M.D., M.P.H.; Donald E. Humphries, Ph.D.; Luis E. Selva, Ph.D.
  VA Boston Healthcare System, 150 S. Huntington Avenue, Boston, MA 02130
- MVP Informatics, Boston – Nhan Do, M.D.; Shahpoor (Alex) Shayan, M.S.
  VA Boston Healthcare System, 150 S. Huntington Avenue, Boston, MA 02130
- MVP Data Operations/Analytics, Boston – Kelly Cho, M.P.H., Ph.D.
  VA Boston Healthcare System, 150 S. Huntington Avenue, Boston, MA 02130
- Director of Regulatory Affairs – Lori Churby, B.S.
  VA Palo Alto Health Care System, 3801 Miranda Avenue, Palo Alto, CA 94304
- MVP Coordinating Centers
  - Cooperative Studies Program Clinical Research Pharmacy Coordinating Center, Albuquerque – Todd Connor, Pharm.D.; Dean P. Argyres, B.S., M.S.
    New Mexico VA Health Care System, 1501 San Pedro Drive SE, Albuquerque, NM 87108
  - Genomics Coordinating Center, Palo Alto – Philip S. Tsao, Ph.D.
    VA Palo Alto Health Care System, 3801 Miranda Avenue, Palo Alto, CA 94304
  - MVP Boston Coordinating Center, Boston - J. Michael Gaziano, M.D., M.P.H.
    VA Boston Healthcare System, 150 S. Huntington Avenue, Boston, MA 02130
  - MVP Information Center, Canandaigua – Brady Stephens, M.S.
    Canandaigua VA Medical Center, 400 Fort Hill Avenue, Canandaigua, NY 14424

**MVP Science**

- Saiju Pyarajan Ph.D.
  VA Boston Healthcare System, 150 S. Huntington Avenue, Boston, MA 02130
- Philip S. Tsao, Ph.D.
  VA Palo Alto Health Care System, 3801 Miranda Avenue, Palo Alto, CA 94304
- Data Core - Kelly Cho, M.P.H, Ph.D.
  VA Boston Healthcare System, 150 S. Huntington Avenue, Boston, MA 02130
- VA Informatics and Computing Infrastructure (VINCI) – Scott L. DuVall, Ph.D.
  VA Salt Lake City Health Care System, 500 Foothill Drive, Salt Lake City, UT 84148
- Data and Computational Sciences – Saiju Pyarajan, Ph.D.
  VA Boston Healthcare System, 150 S. Huntington Avenue, Boston, MA 02130
- Statistical Genetics – Elizabeth Hauser, Ph.D.
  Durham VA Medical Center, 508 Fulton Street, Durham, NC 27705
- Yan Sun, Ph.D.
  Atlanta VA Medical Center, 1670 Clairmont Road, Decatur, GA 30033
- Hongyu Zhao, Ph.D.
  West Haven VA Medical Center, 950 Campbell Avenue, West Haven, CT 06516

**Current MVP Local Site Investigators**

- Atlanta VA Medical Center (Peter Wilson, M.D.)
  1670 Clairmont Road, Decatur, GA 30033
- Bay Pines VA Healthcare System (Rachel McArdle, Ph.D.)
  10,000 Bay Pines Blvd Bay Pines, FL 33744
- Birmingham VA Medical Center (Louis Dellitalia, M.D.)
  700 S. 19th Street, Birmingham AL 35233
- Central Western Massachusetts Healthcare System (Kristin Mattocks, Ph.D., M.P.H.)
  421 North Main Street, Leeds, MA 01053
- Cincinnati VA Medical Center (John Harley, M.D., Ph.D.)
  3200 Vine Street, Cincinnati, OH 45220

- Clement J. Zablocki VA Medical Center (Jeffrey Whittle, M.D., M.P.H.)
  5000 West National Avenue, Milwaukee, WI 53295
- VA Northeast Ohio Healthcare System (Frank Jacono, M.D.)
  10701 East Boulevard, Cleveland, OH 44106
- Durham VA Medical Center (Jean Beckham, Ph.D.)
  508 Fulton Street, Durham, NC 27705
- Edith Nourse Rogers Memorial Veterans Hospital (John Wells., Ph.D.)
  200 Springs Road, Bedford, MA 01730
- Edward Hines, Jr. VA Medical Center (Salvador Gutierrez, M.D.)
  5000 South 5th Avenue, Hines, IL 60141
- Veterans Health Care System of the Ozarks (Kathrina Alexander, M.D.)
  1100 North College Avenue, Fayetteville, AR 72703
- Fargo VA Health Care System (Kimberly Hammer, Ph.D.)
  2101 N. Elm, Fargo, ND 58102
- VA Health Care Upstate New York (James Norton, Ph.D.)
  113 Holland Avenue, Albany, NY 12208
- New Mexico VA Health Care System (Gerardo Villareal, M.D.)
  1501 San Pedro Drive, S.E. Albuquerque, NM 87108
- VA Boston Healthcare System (Scott Kinlay, M.B.B.S., Ph.D.)
  150 S. Huntington Avenue, Boston, MA 02130
- VA Western New York Healthcare System (Junzhe Xu, M.D.)
  3495 Bailey Avenue, Buffalo, NY 14215-1199
- Ralph H. Johnson VA Medical Center (Mark Hamner, M.D.)
  109 Bee Street, Mental Health Research, Charleston, SC 29401
- Columbia VA Health Care System (Roy Mathew, M.D.)
  6439 Garners Ferry Road, Columbia, SC 29209
- VA North Texas Health Care System (Sujata Bhushan, M.D.)
  4500 S. Lancaster Road, Dallas, TX 75216
- Hampton VA Medical Center (Pran Iruvanti, D.O., Ph.D.)
  100 Emancipation Drive, Hampton, VA 23667
- Richmond VA Medical Center (Michael Godschalk, M.D.)
  1201 Broad Rock Blvd., Richmond, VA 23249
- Iowa City VA Health Care System (Zuhair Ballas, M.D.)
  601 Highway 6 West, Iowa City, IA 52246-2208
- Eastern Oklahoma VA Health Care System (River Smith, Ph.D.)
  1011 Honor Heights Drive, Muskogee, OK 74401
- James A. Haley Veterans' Hospital (Stephen Mastorides, M.D.)
  13000 Bruce B. Downs Blvd, Tampa, FL 33612
- James H. Quillen VA Medical Center (Jonathan Moorman, M.D., Ph.D.)
  Corner of Lamont & Veterans Way, Mountain Home, TN 37684
- John D. Dingell VA Medical Center (Saib Gappy, M.D.)
  4646 John R Street, Detroit, MI 48201
- Louisville VA Medical Center (Jon Klein, M.D., Ph.D.)
  800 Zorn Avenue, Louisville, KY 40206
- Manchester VA Medical Center (Nora Ratcliffe, M.D.)
  718 Smyth Road, Manchester, NH 03104
- Miami VA Health Care System (Ana Palacio, M.D., M.P.H.)
  1201 NW 16th Street, 11 GRC, Miami FL 33125
- Michael E. DeBakey VA Medical Center (Olaoluwa Okusaga, M.D.)
  2002 Holcombe Blvd, Houston, TX 77030
- Minneapolis VA Health Care System (Maureen Murdoch, M.D., M.P.H.)
  One Veterans Drive, Minneapolis, MN 55417
- N. FL/S. GA Veterans Health System (Peruvemba Sriram, M.D.)
  1601 SW Archer Road, Gainesville, FL 32608
- Northport VA Medical Center (Shing Shing Yeh, Ph.D., M.D.)
  79 Middleville Road, Northport, NY 11768
- Overton Brooks VA Medical Center (Neeraj Tandon, M.D.)
  510 East Stoner Ave, Shreveport, LA 71101

- Philadelphia VA Medical Center (Darshana Jhala, M.D.)
  3900 Woodland Avenue, Philadelphia, PA 19104
- Phoenix VA Health Care System (Samuel Aguayo, M.D.)
  650 E. Indian School Road, Phoenix, AZ 85012
- Portland VA Medical Center (David Cohen, M.D.)
  3710 SW U.S. Veterans Hospital Road, Portland, OR 97239
- Providence VA Medical Center (Satish Sharma, M.D.)
  830 Chalkstone Avenue, Providence, RI 02908
- Richard Roudebush VA Medical Center (Suthat Liangpunsakul, M.D., M.P.H.)
  1481 West 10th Street, Indianapolis, IN 46202
- Salem VA Medical Center (Kris Ann Oursler, M.D.)
  1970 Roanoke Blvd, Salem, VA 24153
- San Francisco VA Health Care System (Mary Whooley, M.D.)
  4150 Clement Street, San Francisco, CA 94121
- South Texas Veterans Health Care System (Sunil Ahuja, M.D.)
  7400 Merton Minter Boulevard, San Antonio, TX 78229
- Southeast Louisiana Veterans Health Care System (Joseph Constans, Ph.D.)
  2400 Canal Street, New Orleans, LA 70119
- Southern Arizona VA Health Care System (Paul Meyer, M.D., Ph.D.)
  3601 S 6th Avenue, Tucson, AZ 85723
- Sioux Falls VA Health Care System (Jennifer Greco, M.D.)
  2501 W 22nd Street, Sioux Falls, SD 57105
- St. Louis VA Health Care System (Michael Rauchman, M.D.)
  915 North Grand Blvd, St. Louis, MO 63106
- Syracuse VA Medical Center (Richard Servatius, Ph.D.)
  800 Irving Avenue, Syracuse, NY 13210
- VA Eastern Kansas Health Care System (Melinda Gaddy, Ph.D.)
  4101 S 4th Street Trafficway, Leavenworth, KS 66048
- VA Greater Los Angeles Health Care System (Agnes Wallbom, M.D., M.S.)
  11301 Wilshire Blvd, Los Angeles, CA 90073
- VA Long Beach Healthcare System (Timothy Morgan, M.D.)
  5901 East 7th Street Long Beach, CA 90822
- VA Maine Healthcare System (Todd Stapley, D.O.)
  1 VA Center, Augusta, ME 04330
- VA New York Harbor Healthcare System (Peter Liang, M.D., M.P.H.)
  423 East 23rd Street, New York, NY 10010
- VA Pacific Islands Health Care System (Daryl Fujii, Ph.D.)
  459 Patterson Rd, Honolulu, HI 96819
- VA Palo Alto Health Care System (Philip Tsao, Ph.D.)
  3801 Miranda Avenue, Palo Alto, CA 94304-1290
- VA Pittsburgh Health Care System (Patrick Strollo, Jr., M.D.)
  University Drive, Pittsburgh, PA 15240
- VA Puget Sound Health Care System (Edward Boyko, M.D.)
  1660 S. Columbian Way, Seattle, WA 98108-1597
- VA Salt Lake City Health Care System (Jessica Walsh, M.D.)
  500 Foothill Drive, Salt Lake City, UT 84148
- VA San Diego Healthcare System (Samir Gupta, M.D., M.S.C.S.)
  3350 La Jolla Village Drive, San Diego, CA 92161
- VA Sierra Nevada Health Care System (Mostaqul Huq, Pharm.D., Ph.D.)
  975 Kirman Avenue, Reno, NV 89502
- VA Southern Nevada Healthcare System (Joseph Fayad, M.D.)
  6900 North Pecos Road, North Las Vegas, NV 89086
- VA Tennessee Valley Healthcare System (Adriana Hung, M.D., M.P.H.)
  1310 24th Avenue, South Nashville, TN 37212
- Washington DC VA Medical Center (Jack Lichy, M.D., Ph.D.)
  50 Irving St, Washington, D. C. 20422
- W.G. (Bill) Hefner VA Medical Center (Robin Hurley, M.D.)
  1601 Brenner Ave, Salisbury, NC 28144

- White River Junction VA Medical Center (Brooks Robey, M.D.)
  163 Veterans Drive, White River Junction, VT 05009
- William S. Middleton Memorial Veterans Hospital (Prakash Balasubramanian, M.D.)
  2500 Overlook Terrace, Madison, WI 53705

**NTR**

Funding for the Netherland Twin Register was obtained from the Netherlands Organization for Scientific Research (NWO) and The Netherlands Organisation for Health Research and Development (ZonMW) grants 904-61-090, 985-10-002, 904-61-193,480-04-004, 400-05-717, Addiction-31160008, 016-115-035, 400-07-080, Middelgroot-911-09-032, NWO-Groot 480-15-001/674, Center for Medical Systems Biology (CSMB, NWO Genomics), NBIC/BioAssist/RK(2008.024), Biobanking and Biomolecular Resources Research Infrastructure (BBMRI –NL, 184.021.007 and 184.033.111), X-Omics 184-034-019; Spinozapremie (NWO- 56-464-14192), KNAW Academy Professor Award (PAH/6635) and University Research Fellow grant (URF) to DIB; Amsterdam Public Health research institute (former EMGO+) , Neuroscience Amsterdam research institute (former NCA) ; the European Community's Fifth and Seventh Framework Program (FP5- LIFE QUALITY-CT-2002-2006, FP7- HEALTH-F4-2007-2013, grant 01254: GenomEUtwin, grant 01413: ENGAGE); the European Research Council (ERC Starting 284167, ERC Consolidator 771057, ERC Advanced 230374), Rutgers University Cell and DNA Repository (NIMH U24 MH068457-06), the National Institutes of Health (NIH, R01D0042157-01A1, MH081802, DA018673, R01 DK092127-04, Grand Opportunity grant 1RC2 MH089951); the Avera Institute for Human Genetics, Sioux Falls, South Dakota (USA). Part of the genotyping and analyses were funded by the Genetic Association Information Network (GAIN) of the Foundation for the National Institutes of Health.

**SAFS**

Collection of the San Antonio Family Study data was supported in part by National Institutes of Health (NIH) grants P01 HL045522, MH078143, MH078111 and MH083824; and whole genome sequencing of SAFS subjects was supported by U01 DK085524 and R01 HL113323. We are very grateful to the participants of the San Antonio Family Study for their continued involvement in our research programs.

**VIKING**

The Viking Health Study – Shetland (VIKING) was supported by the MRC Human Genetics Unit quinquennial programme grant “QTL in Health and Disease”. DNA extractions and genotyping were performed at the Edinburgh Clinical Research Facility, University of Edinburgh. We would like to acknowledge the invaluable contributions of the research nurses in Shetland, the administrative team in Edinburgh and the people of Shetland.
For the purpose of open access, the author has applied a Creative Commons Attribution (CC BY) license to any Author Accepted Manuscript version arising from this submission.

**WHI**

The WHI program is funded by the National Heart, Lung, and Blood Institute, National Institutes of Health, U.S. Department of Health and Human Services through contracts 75N92021D00001, 75N92021D00002, 75N92021D00003, 75N92021D00004, 75N92021D00005.

#### TOPMed Omics Support Table

| TOPMed Accession # | Parent Study Short Name | TOPMed Phase | Omics Center | Omics Support (WGS) |
|---|---|---|---|---|
| phs000956 | Amish | 1 | Broad Genomics | 3R01HL121007-01S1 |
| phs001211 | ARIC AFGen | 1 | Broad Genomics | 3R01HL092577-06S1 |
| phs001211 | ARIC | 2 | Baylor | 3U54HG003273-12S2 / HHSN268201500015C |
| phs001612 | CARDIA | 3 | Baylor | HHSN268201600033I |
| phs000954 | CFS | 1 | NWGC | 3R01HL098433-05S1 |
| phs000954 | CFS | 3.5 | NWGC | HHSN268201600032I |
| phs001368 | CHS | 3 | Baylor | HHSN268201600033I |
| phs001368 | CHS | 5.5 | Broad Genomics | HHSN268201600034I |
| phs001368 | CHS VTE | 2 | Baylor | 3U54HG003273-12S2 / HHSN268201500015C |
| phs000951 | COPDGene | 1 | NWGC | 3R01HL089856-08S1 |
| phs000951 | COPDGene | 2 | Broad Genomics | HHSN268201500014C |
| phs000951 | COPDGene | 2.5 | Broad Genomics | HHSN268201500014C |
| phs000974 | FHS AFGen | 1 | Broad Genomics | 3R01HL092577-06S1 |
| phs000974 | FHS | 1 | Broad Genomics | 3U54HG003067-12S2 |
| phs000974 | FHS | pilot; 4.5, 5.5, 5.6 | Broad Genomics | HHSN268201600034I |
| phs001218 | GeneSTAR AA_CAC | 2 | Broad Genomics | HHSN268201500014C |
| phs001218 | GeneSTAR | legacy | Illumina | R01HL112064 |
| phs001218 | GeneSTAR | 2 | Psomagen | 3R01HL112064-04S1 |
| phs001345 | GENOA | 2 | NWGC | 3R01HL055673-18S1 |
| phs001345 | GENOA AA_CAC | 2 | Broad Genomics | HHSN268201500014C |
| phs000964 | JHS | 1 | NWGC | HHSN268201100037C |
| phs001416 | MESA AA_CAC | 2 | Broad Genomics | HHSN268201500014C |
| phs001416 | MESA | 2 | Broad Genomics | 3U54HG003067-13S1 |
| phs001416 | MESA | pilot, 5.5, 8 | Broad Genomics | HHSN268201600034I |
| phs001215 | SAFS | 1 | Illumina | 3R01HL113323-03S1 |
| phs001215 | SAFS | legacy | Illumina | R01HL113322 |
| phs001237 | WHI | 2 | Broad Genomics | HHSN268201500014C |

- Baylor = Baylor College of Medicine Human Genome Sequencing Center
- Broad Genomics = Broad Institute Genomics Platform
- Illumina = Illumina
- NWGC = Northwest Genomics Center
- Psomagen = Psomagen

### Supplemental Methods

#### 1. Study Descriptions

**Airwave**

The Airwave Health Monitoring Study was launched in 2004 with the aim to recruit over 60,000 participants from police forces in Great Britain by 2018. The study includes two phases. First, every employee receives the enrolment questionnaire via routine administration or the occupational health service. The second phase is a health screen performed locally by trained nurses using a standardised protocol (Fig. 1). Participants can attend the health screen irrespective of their participation in phase 1 and their Airwave usage. In phase 2, volunteers are recruited through general force-wide publicity (emails, wall posters, and articles in newsletters), word of mouth or direct contact if they request a health screen on their enrolment questionnaire. Blood and urine samples are obtained at the screening visit (with consent) and stored for future research use. The study has ethical approval from the National Health Service Multi-site Research Ethics Committee (MREC/13/NW/0588)[1].

**AMISH**

The Amish Research Program, based at the University of Maryland, Baltimore, includes a set of large community-based studies focused largely on cardiometabolic health carried out in the Old Order Amish (OOA) community of Lancaster County, Pennsylvania. Over 9,000 Amish have been recruited since 1995 into one or more protocols. The Lancaster Amish community is a founder population who immigrated to Pennsylvania from Western Europe in the early 1700’s, later expanding into other regions of the U.S. The Amish cohort participating in the TOPMed Consortium comprises 1,120 subjects ≥ 18 years of age from large multigenerational families who were recruited for specific protocols between 2001 and 2006. Subjects have been extensively phenotyped for a range of cardiometabolic traits, including anthropometry, lipids, blood pressure, glucose and related measures, vascular imaging, and a range of other phenotypes. DNA samples have been collected and serum and plasma samples biobanked.

**ARIC**

The Atherosclerosis Risk in Communities (ARIC) study recruited 15,792 adults aged 45 to 64 years in 1987 through 1989 by probability sampling from Forsyth County, North Carolina; Jackson, Mississippi; suburbs of Minneapolis, Minnesota; and Washington County, Maryland[2]. The Jackson sample comprised African Americans only; the other three samples represent the ethnic mix of their communities. Extensive information was collected at baseline on cardiovascular risk factors. The ARIC study was approved by the institutional review board of each field center institute and participants gave informed consent including consent for genetic testing.

**CAERPHILLY**

The Caerphilly Prospective study (CaPs) was a cohort study of men, initially established to look at cardiovascular disease. The electoral roll and general practice lists were used to identify eligible participants (men aged 45 to 59 years) from the town of Caerphilly and adjoining villages in South Wales, United Kingdom. Participation rate was 89% and 2,512 men were examined in phase one from July 1979 until September 1983. Men were followed up around every 4 to 5 years for cardiovascular outcomes and later for dementia cognitive impairment and aging traits. Written informed consent was obtained from all participants. Approval for the original CaPS was obtained from the Research Ethics Committee in South Glamorgan, Wales. Approval for the phase five follow-up study was obtained from the Research Ethics Committee in Gwent, Wales.

**CARDIA**

The Coronary Artery Risk Development in Young Adults (CARDIA) Study is a prospective multicenter study with 5115 Caucasian and African American participants ages 18-30 years at baseline, recruited from four centers. The recruitment was done from the total community in Birmingham, AL, from selected census tracts in Chicago, IL and Minneapolis, MN; and from the Kaiser Permanente health plan membership in Oakland, CA. The details of the study design for the CARDIA study have been published before[3]. Nine examinations have been completed since the baseline examination in 1985–1986, with follow-up examinations 2, 5, 7, 10, 15, 20, 25, 30 years after baseline.

Written informed consent was obtained from participants at each examination and all study protocols were approved by the institutional review boards of the participating institutions.

**CFS**
The Cleveland Family Study (CFS) was designed to examine the genetic basis of sleep apnea in 2,534 African-American and European-American individuals from 356 families. Index probands with confirmed sleep apnea were recruited from sleep centers in northern Ohio, supplemented with additional family members and neighborhood control families [{Redline1995}]. Four visits occurred between 1990 and 2006; in the first 3, data were collected in participants' homes while the last occurred in a clinical research center (2000 - 2006). Measurements included sleep apnea monitoring, blood pressure, anthropometry, spirometry and other related phenotypes. Blood samples (overnight fasting, before bed and following an oral glucose tolerance test), nasal and oral ultrasound, and ECG were also obtained during the 4th exam. Institutional Review Board approval and signed informed consent was obtained for all participants.

**CHRIS**
The CHRIS study is a prospective study set up in 2011 in the Val Venosta/Vinschgau district, South Tyrol, Italy. Recruited into the study at baseline between 2011 and 2018 were 13,393 participants from 13 municipalities, each one characterized by a central town, small villages, and scattered mountain farms. Settlements are located at an altitude of 600 to 2000 m above sea level. Participants cover more than one-third of the target region population. A full description of the study has been previously published[4].

**CHS**
The Cardiovascular Health Study (CHS) is a population-based cohort study of risk factors for CHD and stroke in adults ≥65 years conducted across 4 field centers[5]. The original predominantly Caucasian cohort of 5,201 persons was recruited in 1989-1990 from random samples of the Medicare eligibility lists; subsequently, an additional predominantly African American cohort of 687 persons was enrolled in 1992-1993 for a total sample of 5,888. DNA was extracted from blood samples drawn on all participants at their baseline examination and all participants provided informed consent for the use of their genetic data in analyses. After an 8-12-h fast, CHS participants underwent phlebotomy by atraumatic venipuncture with a 21-gauge butterfly needle connected to a Vacutainer (Becton Dickinson, Rutherford, NJ) outlet via a Luer adaptor[6].

**COPDGene**
COPDGene (ClinicalTrials.gov: NCT00608764) is an ongoing study of over 10,000 non-Hispanic White and African American cigarette smokers. It was designed to investigate COPD and other smoking-related lung diseases[7]. As part of TOPMed freeze 6a, WGS was conducted on 10,372 subjects.

**CROATIA studies**
The **CROATIA-Split** study is a population-based, cross-sectional study in the Dalmatian City of Split in Croatia that includes 1000 examinees aged 18-95. Blood samples were collected in 2009 and 2010 along with many clinical and biochemical measures and lifestyle and health questionnaires. A detailed description of the study has been published elsewhere[8].

The **CROATIA-Korcula** study is a family-based, cross-sectional study in the isolated island of Korcula in Croatia that included 965 examinees aged 18-95. Blood samples were collected in 2007 along with many clinical and biochemical measures and lifestyle and health questionnaires. A detailed description of the study has been published elsewhere[9].

The **CROATIA-Vis** study is a family-based, cross-sectional study in the isolated island of Vis in Croatia that included 1,056 examinees aged 18-93. Blood samples were collected in 2003 and 2004 along with many clinical and biochemical measures and lifestyle and health questionnaires. A detailed description of the study has been published elsewhere[10].

**EPIC-Norfolk**

The European Prospective Investigation in Cancer (EPIC) Norfolk Study (EPIC-Norfolk) The EPIC-Norfolk GWAS consists of 1,284 obese individuals and 2,566 random subcohort in a case-cohort design randomly selected from the EPIC Norfolk Study, a population-based cohort study of 25,663 men and women of European descent aged 39-79 years recruited in Norfolk, UK between 1993 and 1995[11]. Height and weight were measured using standard anthropometric techniques. After quality controls, 3,552 individuals remained in the final sample. The Norwich Local Research Ethics Committee granted ethical approval for the study. All participants gave written informed consent.

**FHS**

The Framingham Heart Study (FHS) was started in 1948 with 5,209 randomly ascertained participants from Framingham, Massachusetts, US, who had undergone biannual examinations to investigate cardiovascular disease and its risk factors. In 1971, the Offspring cohort (comprising 5,124 children of the original cohort and the children's spouses) and in 2002, the Third Generation (consisting of 4,095 children of the Offspring cohort) were recruited. FHS participants in this study are of European ancestry. The methods of recruitment and data collection for the Offspring and Third Generation cohorts have been described[12].

**GAIT2**

The Genetic Analysis of Idiopathic Thrombophilia(GAIT) project is a family based study where 935 subjects in 35 extended pedigrees were collected [13,14]. To be included in the study, a family was required to have at least 10 living individuals in 3 or more generations. Families were selected through a proband with idiopathic thrombophilia, which was defined as recurrent thrombotic events (at least one of which was spontaneous), a single spontaneous thrombotic episode plus a first-degree relative also affected, or onset of thrombosis before age 45. Thrombosis in these probands was considered idiopathic when biological causes as antithrombin deficiency, protein S and C deficiencies, activated protein C resistance, plasminogen deficiency, heparin cofactor II deficiency, Factor V Leiden, dysfibrinogenemia, lupus anticoagulant and antiphospholipid antibodies, were excluded. Subjects were interviewed by a physician to determine their health and reproductive history, current medications, alcohol consumption, use of sex hormones (oral contraceptives or hormonal replacement therapy) and their smoking history. The study was performed according to the Declaration of Helsinki. All procedures of the study were reviewed by the Institutional Review Board of the Hospital de la Santa Creu i Sant Pau, Barcelona, Spain. Adult subjects gave informed consent for themselves and for their minor children.

**GeneSTAR**

GeneSTAR (Genetic Study of Atherosclerosis Risk) is an ongoing prospective family study begun in 1983 to determine environmental, phenotypic, and genetic causes of premature cardiovascular disease[15]. Briefly, probands with a premature coronary disease event prior to 60 years of age were identified at the time of hospitalization in any of 10 Baltimore area hospitals. Their apparently healthy 30-59 year old siblings without known CAD were recruited and underwent phenotypic measurement and characterization between 1983 and 2006; offspring of the siblings and probands, as well as the co-parent of these offspring, were recruited and assessed between 2003 and 2006. All participants provided written informed consent. Participants are followed for coronary artery disease events at regular intervals.

**GENOA**

The Genetic Epidemiology Network of Arteriopathy (GENOA) study is one of four networks in the NHLBI Family-Blood Pressure Program (FBPP)[16]. GENOA's long-term objective is to elucidate the genetics of target organ complications of hypertension, including both atherosclerotic and arteriolosclerotic complications involving the heart, brain, kidneys, and peripheral arteries. The longitudinal GENOA Study recruited European-American and African-American sibships with at least two individuals with clinically diagnosed essential hypertension before age 60 years. All other members of the sibship were invited to participate regardless of their hypertension status. Exclusion criteria were secondary hypertension, alcoholism or drug abuse, pregnancy, insulin-dependent diabetes mellitus, or active malignancy. Study protocols were approved by the University of Michigan and Mayo Clinic Institutional Review Boards and participants gave written informed consent.

**GOYA**

The GOYA (Male) cohort is a longitudinal case-cohort (obese, non-obese) study comprising a randomly (1%) selected control group and all extremely overweight men identified among 362,200 Caucasian men examined at the mean age of 20 years at the draft boards in Copenhagen and its surrounding areas during 1943–1977. Obesity was defined as 35% overweight relative to a local standard in use at the time (mid 1970's), corresponding to a BMI ≥31.0 kg/m2, which proved to be above the 99th percentile. All of the obese and 50% of the random sampled controls, who were still living in the region, were invited to a follow-up survey in 1992–94 at the mean age of 46 years, at which time the blood samples were taken and genotyping were performed for a total of 673 extremely overweight and 792 controls.(32) With a sampling fraction of 0.5% (50% of 1%), the controls represent about 158,000 men among whom the case group was the most obese.

**HBCS**

The Helsinki Birth Cohort Study (HBCS) is composed of 8 760 individuals born between the years 1934-44 in one of the two main maternity hospitals in Helsinki, Finland. Between 2001 and 2003, a randomly selected sample of 928 males and 1 075 females participated in a clinical follow-up study with a focus on cardiovascular, metabolic and reproductive health, cognitive function and depressive symptoms. Detailed information on the selection of the HBCS participants and on the study design can be found elsewhere[17–19]. Research plan of the HBCS was approved by the Institutional Review Board of the National Public Health Institute and all participants have signed an informed consent.

**INTER99**

The Inter99 is a population-based cohort from 11 municipalities in the south-western part of Copenhagen. This population has been described in detail previously[20]. In brief, the Inter99 study is a randomized, non-pharmacological intervention study for prevention of ischemic heart disease on more than 13,000 individuals between 30 and 60 years randomly selected from the Civil Registration System. Out of those, 6,784 participants attended the baseline health examination, of whom 6,127 had genotype information available. The study was conducted at the Research Centre for Prevention and Health in Glostrup, Denmark.

**JHS**

The Jackson Heart Study (JHS) is a longitudinal investigation of the genetic and environmental risk factors associated with cardiovascular disease in African Americans. JHS recruited 5306 African American residents living in the Jackson, Mississippi, metropolitan area of Hinds, Madison, and Rankin Counties from 2000-2004[21,22]. The age at enrollment for the unrelated cohort was 35-84 years; the nested family cohort (1,498 members of 264 families) included related individuals >21 years old. Participants provided extensive medical, family, lifestyle, and psychosocial histories, had an array of physical and biochemical measurements and diagnostic procedures, and provided biospecimens for future research at baseline and two follow-up examinations (2005 –2008, and 2009 – 2013), with a third follow-up examination in progress[23,24]. Annual follow-up interviews and cohort surveillance for cardiovascular events and mortality are also ongoing[25].

**KORA**

The Monitoring of Trends and Determinants in Cardiovascular Disease/Cooperative Health Research in the Region of Augsburg Study (KORA) consisted of a series of independent population-based epidemiological surveys of participants living in the region of Augsburg, Southern Germany[26,27]. All survey participants are residents of German nationality identified through the registration office and underwent standardized examinations including blood withdrawals for plasma and DNA. The presented data were derived from the KORA surveys S4 (conducted in 1999-2001) and F4 (conducted in 2006-2008) and comprised 3,720 participants with available fibrinogen and DNA information.

**LBC studies**

The Lothian Birth Cohort (LBC) studies, LBC1936 & LBC1921, were ascertained as follows. The **LBC1936** consists of 1,091 relatively healthy individuals assessed on cognitive and medical traits at 70 years of age. They were born in 1936, most took part in the Scottish Mental Survey of 1947, and almost all lived independently in the Lothian region of Scotland (Edinburgh City and surrounding area). A full description of participant recruitment and testing can be found elsewhere[28,29]. The **LBC1921** cohort consists of 550 relatively healthy individuals, 316 females and

234 males, assessed on cognitive and medical traits at 79 years of age. They were born in 1921, most took part in the Scottish Mental Survey of 1932, and almost all lived independently in the Lothian region in Scotland. A full description of participant recruitment and testing can be found elsewhere[28,30]. Ethics permission for the study was obtained from the Multi-Centre Research Ethics Committee for Scotland (MREC/01/0/56) and from Lothian Research Ethics Committee (LBC1936: LREC/2003/2/29 and LBC1921: LREC/1998/4/183). The research was carried out in compliance with the Helsinki Declaration. All subjects gave written, informed consent.

**LURIC**

The Ludwigshafen Risk and Cardiovascular Health (LURIC) study is an ongoing prospective study of more than 3,300 individuals of German ancestry in whom cardiovascular and metabolic phenotypes (CAD, MI, dyslipidemia, hypertension, metabolic syndrome and diabetes mellitus) have been defined or ruled out using standardized methodologies in all study participants. Inclusion criteria for LURIC were: German ancestry (limitation of genetic heterogeneity), clinical stability (except for acute coronary syndromes) and availability of a coronary angiogram. Exclusion criteria were: any acute illness other than acute coronary syndromes, any chronic disease where non-cardiac disease predominated and a history of malignancy within the last five years. Genome-wide analyses using the Affymetrix 6.0 have been completed in all participants. A 10-year clinical follow-up for total and cause specific mortality has been completed.

**MARTHA**

The MARseille THrombosis Association (MARTHA) project has been previously described[31]. Briefly, MARTHA consist in two independent samples of VT patients, named MARTHA08 (N=1,006) and MARTHA10 (N=586). MARTHA patients are unrelated subjects of European origin, with the majority being of French ancestry, consecutively recruited at the Thrombophilia center of La Timone hospital (Marseille, France) between January 1994 and October 2005. All patients had a documented history of VT and free of well characterized genetic risk factors including AT, PC, or PS deficiency, homozygosity for FV Leiden or FII 20210A, and lupus anticoagulant. They were interviewed by a physician on their medical history, which emphasized manifestations of deep vein thrombosis and pulmonary embolism using a standardized questionnaire. The thrombotic events were confirmed by venography, Doppler ultrasound, spiral computed tomographic scanning angiography, and/or ventilation/perfusion lung scan.

**MESA**

The Multi-Ethnic Study of Atherosclerosis (MESA) is a cohort study designed to investigate the characteristics of subclinical cardiovascular disease and the risk factors that predict progression to clinically overt cardiovascular disease or progression of the subclinical disease. MESA comprises a diverse, population-based sample of 6,814 asymptomatic men and women aged 45-84. Thirty-eight percent of the recruited participants are Caucasian, 28 percent African-American, 22 percent Hispanic, and 12 percent Asian, predominantly of Chinese descent[32]. Participants were recruited from six field centers across the United States: Wake Forest University, Columbia University, Johns Hopkins University, University of Minnesota, Northwestern University and University of California - Los Angeles. In this study we included only European American participants.

**MVP**

The Veterans Affairs Million Veteran Program (MVP) started recruiting US military veterans from 63 Veterans Affairs (VA) facilities across the United States in 2011. Veterans aged 18 years and older are recruited into MVP where participants are linked to VA electronic health records (EHR), complete a questionnaire, and submit a blood sample at enrollment. The EHR includes information on inpatient International Classification of Disease (ICD) diagnosis codes, Current Procedural Terminology (CPT) procedure codes, and clinical laboratory measurements[33].

**NEO**

The NEO was designed for extensive phenotyping to investigate pathways that lead to obesity-related diseases. The NEO study is a population-based, prospective cohort study that includes 6,671 individuals aged 45–65 years, with an oversampling of individuals with overweight or obesity. At baseline, information on demography, lifestyle, and medical history have been collected by questionnaires. In addition, samples of 24-h urine, fasting and postprandial blood plasma and serum, and DNA were collected. Genotyping was performed using the Illumina HumanCore Exome chip, which was subsequently imputed to the 1000 genome reference panel. Participants underwent an extensive physical examination, including anthropometry, electrocardiography, spirometry, and measurement of the carotid artery intima-media thickness by ultrasonography. In random subsamples of participants, magnetic resonance imaging of abdominal fat, pulse wave velocity of the aorta, heart, and brain, magnetic resonance spectroscopy of the liver, indirect calorimetry, dual energy X-ray absorptiometry, or accelerometry measurements were performed. The collection of data started in September 2008 and completed at the end of September 2012. Participants are currently being followed for the incidence of obesity-related diseases and mortality.

**NTR**

As part of a Netherlands Twin Registry (NTR) biobank project, 9,530 participants from 3,477 families were visited at home between January 2004 and July 2008 for collection of blood samples. Visits were scheduled between 7:00 and 10:00 am and fertile women were bled on day 2–4 of the menstrual cycle, or in their pill-free week. Fertile women were bled on day 2–4 of the menstrual cycle, or in their pill-free week. Body composition was measured and information about physical health and lifestyle (e.g. smoking and drinking behavior, physical exercise, medication use) was obtained. For more detailed information about the methodology of the NTR Biobank study, see[34]. The NTR studies were approved by the Central Ethics Committee on Research involving human subjects of the VU University Medical Center, Amsterdam, an Institutional Review Board certified by the US Office of Human Research Protections (IRB number IRB-2991 under Federal wide Assurance-3703; IRB/institute codes, NTR 03-180). All subjects provided written informed consent. Valid GWA data were available for 6171 individuals.

**PROCARDIS**

The Precocious Coronary Artery Disease Study (PROCARDIS) consists of coronary artery disease (CAD) cases and controls from four European countries (UK, Italy, Sweden and Germany). CAD (defined as myocardial infarction, acute coronary syndrome, unstable or stable angina, or need for coronary artery bypass surgery or percutaneous coronary intervention) was diagnosed before 66 years of age and 80% of cases had a sibling fulfilling the same criteria for CAD. Subjects with self-reported non-European ancestry were excluded. Among the “genetically-enriched” CAD cases, 70% had suffered myocardial infarction (MI).

**PROSPER**

PROspective Study of Pravastatin in the Elderly at Risk (PROSPER) was a prospective multicenter randomized placebo-controlled trial to assess whether treatment with pravastatin diminishes the risk of major vascular events in elderly[35,36]. Between December 1997 and May 1999, we screened and enrolled subjects in Scotland (Glasgow), Ireland (Cork), and the Netherlands (Leiden). Men and women aged 70-82 years were recruited if they had pre-existing vascular disease or increased risk of such disease because of smoking, hypertension, or diabetes. A total number of 5804 subjects were randomly assigned to pravastatin or placebo. A large number of prospective tests were performed including Biobank tests and cognitive function measurements. A detailed description of the study has been published elsewhere[35,37].

**RETROVE**

RETROVE is a prospective case–control study that includes 400 consecutive patients with VTE (cancer associated thrombosis was excluded) and 400 healthy control volunteers[38]. All individuals were ≥ 18 years. The diagnosis was confirmed with Doppler ultrasonography, tomography, magnetic resonance, arteriography, phlebography or pulmonary gammagraphy. Blood samples from the patients were taken at least 6 months after thrombosis to minimize the influence of the acute phase. None of the participants was using oral anticoagulants, heparin, or antiplatelet therapy at the time of blood collection. Controls were selected according to the age and sex distribution of the Spanish population (2001 census). A total of 5 ml of blood was obtained in a Vacutainer tube (BD Vacutainer Becton Dickinson and Company, New Jersey, USA) containing EDTA as anticoagulant. All individuals were genotyped using Infinium Global Screening Array-24 v3.0 kit from Illumina. Written informed consent was obtained for all participants and all procedures were approved by the Institutional Review Board of the Hospital de la Santa Creu i Sant Pau (Barcelona).

**RS**

The Rotterdam Study (RS-I and RS-II) is a prospective, population-based cohort study of determinants of several chronic diseases in older adults[39]. RS-I comprised 7,983 inhabitants of Ommoord, a district of Rotterdam in the Netherlands, who were 55 years or over. The baseline examination took place between 1990 and 1993. In 1999, the cohort was extended to include 3011 inhabitants who reached the age of 55 years after the baseline examination and persons aged 55 years or older who migrated into the research area (RS-II). Subjects are of European ancestry based on their self-report.

**SardiNIA**

The SardiNIA study has been previously described[40]. Briefly, it is a large population-based study which consists of 6,921 individuals, males and females, ages 14-102 y, and representing >60% of the adult population of four villages in the Lanusei Valley of Sardinia. Samples have been characterized for several quantitative traits and medical conditions, including fibrinogen.

**SAFS**

San Antonio Family Study (SAFS): The SAFS is a longitudinal study which began in 1991 and was designed to primarily investigate the genetics of cardiovascular disease and its risk factors in Mexican Americans. For this study, subjects from the SAFS included 1,431 individuals in 42 extended families at baseline, from the San Antonio Family Heart Study[41]. Ascertainment occurred by way of the random selection of an adult Mexican American proband, without regard to presence or absence of disease.

**SHIP**

The Study of Health in Pomerania (SHIP) is a longitudinal cohort study in West Pomerania, the north-east area of Germany and has been described previously[42,43]. From the entire study population of 212,157 inhabitants living in the area, a sample was selected from the population registration offices, where all German inhabitants are registered. Only individuals with German citizenship and main residency in the study area were included. A two-stage cluster sampling method was adopted from the WHO MONICA Project Augsburg, Germany. In a first step, the three cities of the region (with 17,076 to 65,977 inhabitants) and the 12 towns (with 1,516 to 3,044 inhabitants) were selected. Further 17 out of 97 smaller towns (with less than 1,500 inhabitants) were drawn at random. In a second step, from each of the selected communities, subjects were drawn at random, proportional to the population size of each community and stratified by age and gender. Finally, 7,008 subjects aged 20 to 79 years were sampled, with 292 persons of each gender in each of the twelve five-year age strata. In order to minimize drop-outs by migration or death, subjects were selected in two waves. The net sample (without migrated or deceased persons) comprised 6,267 eligible subjects. The SHIP population finally comprised 4,308 participants at baseline (corresponding to a final response of 68.8%).

**TwinsUK**

The TwinsUK cohort was derived from the UK adult twin registry based at King's College London (www.twinsUK.ac.uk). These unselected twins have been recruited from the general population through national media campaigns in the United Kingdom and shown to be comparable to age-matched population singletons in terms of disease-related and lifestyle characteristics[44]. Informed consent was obtained from all participants and the study was approved by the St. Thomas' Hospital Ethics Committee.

**VIKING**

The Viking Health Study - Shetland (VIKING) is a family-based, cross-sectional study that seeks to identify genetic factors influencing cardiovascular and other disease risk in the population isolate of the Shetland Isles in northern Scotland[45]. 2,105 participants were recruited between 2013 and 2015, most having at least three grandparents from Shetland. Fasting blood samples were collected and many health-related phenotypes and environmental exposures were measured in each individual. Common and rare genetic variants reveal the gene pool to be distinct from the rest of the British Isles[46,47], consistent with the high levels of endogamy historically. All participants gave informed consent and the study was approved by the South East Scotland Research Ethics Committee, NHS Lothian (reference: 12/SS/0151).

**WGHS**

The Women's Genome Health Study (WGHS) is a prospective cohort of initially healthy, female North American health care professionals at least 45 years old at baseline representing participants in the Women's Health Study (WHS) who provided a blood sample at baseline and consent for blood-based analyses[48]. The WHS was a 2x2 trial beginning in 1992-1994 of vitamin E and low dose aspirin in prevention of cancer and cardiovascular disease with about 10 years of follow-up. Since the end of the trial, follow-up has continued in observational mode. Additional information related to health and lifestyle were collected by questionnaire throughout the WHS trial and continuing observational follow-up.

**WHI**

The Women's Health Initiative (WHI) is one of the largest (n=161,808) studies of women's health ever undertaken in the U.S.(57) There are two major components of WHI: (1) a Clinical Trial (CT) that enrolled and randomized 68,132 women ages 50 – 79 into at least one of three placebo-control clinical trials (hormone therapy, dietary modification, and calcium/vitamin D); and (2) an Observational Study (OS) that enrolled 93,676 women of the same age range into a parallel prospective cohort study. DNA was extracted by the Specimen Processing Laboratory at the Fred Hutchinson Cancer research Center (FHCRC) using specimens that were collected at the time of enrollment.

#### 2. Fibrinogen Measurement

**Airwave**

Blood and urine samples for the Airwave Health Monitoring Study were obtained at the screening visit (with consent) and stored for future research use. Blood samples are spun at the clinics, stored in a thermoporter (Laminar Medica) and sent overnight from the clinics by courier to a dedicated laboratory for analysis. The blood samples are either analysed next day for haematology, coagulation and biochemistry tests or frozen for long-term storage. Blood and urine samples are stored at −80 °C at the laboratory, before being transferred to a bio-repository facility at Hammersmith Hospital (London, United Kingdom) which is the primary long-term storage location for the samples. The samples are stored in vapour phase liquid nitrogen allowing long-term access for biochemical or genetic analysis. Samples for which consent has not been obtained for long-term storage are destroyed. All samples are stored in barcoded tubes. Fibrinogen was measured from sodium citrate plasma using either ACL 300 or 8000 analyser (Beckman Coulter)[1].

**AMISH**

Fibrinogen was measured using the Clauss method[49].

**ARIC**

Fibrinogen was measured at baseline in the entire ARIC cohort after an 8-hour fasting period. Circulating plasma fibrinogen was measured by the Clauss clotting rate method[49]. Participants whose fibrinogen measurement was off 6SD from the mean were also excluded.

**CAERPHILLY**

Blood was taken between 7 and 10 AM for 91% of the men. The blood was collected without venous stasis into evacuated containers with a 19-gauge butterfly needle and Sarstedt monovette adaptors. Centrifugation was performed within 1 hour. Citrated plasma stored at 70°C was used for all samples assayed in the Department of Medicine, University of Glasgow, and fresh plasma samples anticoagulated with edetic acid (EDTA) were used for the measurement of nephelometric fibrinogen (Department of Hematology, Southmead Hospital, Bristol). Two assays for fibrinogen were used - Clauss and heat-nephelometry[50].

**CARDIA**

Blood used for fibrinogen measurement was collected at the baseline exam. Participants were asked to fast for 12 hours prior to blood draw. CARDIA measured fibrinogen by immunonephelometry.

**CFS**
Fibrinogen concentrations were quantified by the STa-R automated coagulation analyzer (Diagnostica Stago, Parsippany, NJ), which uses the clotting method developed by Clauss[49] in which the level of fibrinogen is directly correlated with the clotting time of a diluted plasma sample in the presence of excess thrombin.

**CHRIS**
After overnight fasting, blood samples were collected between 08:00 AM and 10:00 AM at the study center. After pre-analytical sample processing, samples were shipped to the laboratory of the Merano hospital, Italy. There, two different systems were used to assess hemostatic factors using the Clauss method[49] in an aliquot of citrated plasma: 1) ROCHE CA1500 system (between August 2011 and January 2014) and 2) Stago STA Compact max system (between February 2014 and December 2018).

**CHS**
After an 8-12-h fast, CHS participants underwent phlebotomy by atraumatic venipuncture with a 21-gauge butterfly needle connected to a Vacutainer (Becton Dickinson, Rutherford, NJ) outlet via a Luer adaptor[6]. For fibrinogen determination, an additional citrate-containing tube was processed at 4°C. The study measured fibrinogen levels using the Clauss method[49].

**COPDGene**
Peripheral venous blood was collected into Vacutainer tubes, in the morning, after fasting overnight, at baseline and at the one year follow-up visit. Plasma (EDTA as the anticoagulant) was obtained by centrifugation at 2000 g for 10 to 15 minutes. Samples were stored at –80° until analyzed centrally. Fibrinogen (K-ASSAY fibrinogen test, Kamiya Biomedical Co., Seattle, WA, USA) levels were measured using immunoturbidometric assays validated for use with EDTA plasma. The lower limit of quantification (LLQ) for fibrinogen was 5.4 mg/dL, respectively[51].

**CROATIA studies**
The CROATIA studies used the Clauss method for measuring plasma fibrinogen[49].

**EPIC-Norfolk**
Fibrinogen concentration in g/L: A non-fasting blood sample (42 mL) was collected at baseline health check in 1993-1997. Samples later used for fibrinogen analyses were collected in citrated bottles, stored in refrigerator at 4°C overnight and transferred the following morning to the EPIC laboratory for processing. Plasma not required for immediate analysis was frozen in aliquots in liquid nitrogen at -196°C. Between 2000 and 2002 aliquots of plasma were retrieved from liquid nitrogen and thawed for fibrinogen analysis. Fibrinogen was measured by a functional assay based on the method of Clauss using the commercial kit Fibriquik (bioMerieux, Lyon, France) on an MDA180 automated analyser (bioMerieux)[49,52].

**FHS &FHS-omni**
Plasma fibrinogen levels were measured using the Clauss method[49] for both FHS and FHS-omni.

**GAIT2**
Fibrinogen was measured by the Clauss method with thrombin from Diagnostica Stago (Ansières, France)[49].

**GeneSTAR**
After an overnight fast, blood was obtained from venipuncture and collected into a vacutainer tube containing 3.2% sodium citrate after the first 4 mL was discarded. Plasma fibrinogen was measured on an automated optical clot detection device (Behring Coagulation System; Dade-Behring, Newark, Del) using a modified Clauss method. Excess thrombin was added to the citrated plasma, and the time required for clot formation was recorded in seconds; clotting time was converted to mg/dL of fibrinogen by extrapolating the results from a standard fibrinogen curve.

**GENOA**
Blood was drawn after an overnight fast. Fibrinogen was measured from citrated blood by the Clauss (clotting time-based) method[49].

**GOYA**
A functional photometric assay was employed to estimate fibrinogen concentration. The sample is mixed with a snake venom enzyme (Batroxobin) and fibrin formation is recorded turbidimetrically at 334 nm. Reaction conditions are such that a linear increase in absorbance is obtained over a concentration range of fibrinogen from 80-700 mg/dl. Higher or lower ranges can be measured by adjusting the sample volume. Calibration is performed with a single standard[53].

**HBCS**
Fibrinogen levels were measured using the Clauss method with an electrical impedance end point[49].

**INTER99**
In the Inter99 study baseline fasting serum samples were collected and an ultrasensitive immunoassay was used to measure various candidate biomarkers including fibrinogen. Details regarding this assay have been described previously[54].

**JHS**
Fibrinogen antigen assays were performed using a Siemens BNII nephelometer (Erlangen, Germany).

**LBC studies**
Fibrinogen levels were measured using HemosILTM based on the Clauss method[49].

**LURIC**
Fasting blood samples were collected at baseline in the morning before angiography. Fibrinogen was measured in citrate plasma using the Clauss method (STA fibrinogen/STA Stago, Stago Diagnostica/Roche Mannheim, Germany) at the Haemostaseology Laboratory of the Ludwigshafen hospital on a daily basis[49].

**MARTHA**
Blood samples were collected by antecubital venipuncture into Vacutainer® tubes 0.105 M trisodium citrate (ratio 9:1, Becton Dickinson) for the coagulation test and the thrombin generation assay. Platelet-poor plasma (PPP) was obtained after double centrifugation of citrated blood (3000 g for 10 min at 25°C) and kept frozen at -80°C until analysis. Fibrinogen levels were measured using the Clauss method on STAR automatic coagulomater[49].

**MESA**
Fasting blood samples were collected, processed, and stored using standardized procedures. Fibrinogen antigen was measured using the BNII nephelometer (N Antiserum to Human Fibrinogen; Dade Behring Inc., Deerfield, IL). The assay was performed at the Laboratory for Clinical Biochemistry Research (University of Vermont, Burlington, VT). Intra- and inter-assay analytical coefficients of variation were 2.7% and 2.6%, respectively.

**NEO**
After 5 minutes rest, fasting blood samples were collected from the antecubital vein. Serum, heparin-plasma, citrated-plasma, and EDTA-plasma were collected from the blood samples. Plasma was obtained by centrifugation at 2500 g for 10 minutes at room temperature and stored in aliquots at −80°C until testing. Blood samples for measurements of coagulation factors activity were drawn into tubes containing 0.106M trisodium citrate (Sarstedt). Fasting fibrinogen levels were measured according to the method of Clauss[49].

**NTR**
Fibrinogen was measured in a 4.5 ml CTAD tube that was stored during transport in melting ice and upon arrival at the laboratory, centrifuged for 20 minutes at 2000x g at 4° C, after which citrated plasma was harvested, aliquoted

(0.5 ml), snapfrozen in dry ice, and stored at –30° C. Fibrinogen levels were determined using the Clauss method[49] on a STA Compact Analyzer Diagnostica Stago, France), using STA Fibrinogen (Diagnostica Stago, France).

**PROCARDIS**

Blood samples were drawn between 8.00 and 11.00 a.m.at the time of recruitment of cases and controls. Plasma fibrinogen concentrations for the Procardis-clauss sub-sample were measured in fasting citrate plasma samples by the Clauss method using the IL Test Fibrinogen C kit and IL Test Calibration Plasma, on the ACL-9000 coagulometer (all from Instrumentation Laboratory Spa, Milan, Italy)[49]. The inter-assay CV was 7% (n=106). For the Procardis-immunonephelometric, fibrinogen was measured in EDTA plasma samples using Dade Behring reagents on the Dade-Behring Nephelometer II analyzer (Dade-Behring, Marburg, Germany). The inter-assay CV was 5.5%.

**PROSPER**

Fibrinogen levels were measured by the Clauss method using aMDA180 coagulometer (Trinity Biotech; calibrant 9th British standard National Institute for Biological Standards and Control)[49].

**RETROVE**

A total of 5 ml of blood was obtained in a Vacutainer tube (BD Vacutainer Becton Dickinson and Company, New Jersey, USA) containing citrate as anticoagulant. Fibrinogen was measured by the Clauss method with thrombin from Diagnostica Stago (Ansières, France) and expressed in g/l[49].

**RS**

In RS-I, fibrinogen levels were derived at baseline (RS-I-1) from the clotting curve of the prothrombin time assay using Thromborel S as a reagent on an automated coagulation laboratory 300 (ACL 300, Instrumentation Laboratory, Zaventem, Belgium). At the second follow up of RS-I (RS-I-3) and the baseline visit of RS-II, fibrinogen levels were derived from the clotting curve of the prothrombin time assay using Thromborel S (Behringwerke, Marburg, Germany) as a reagent on an automated coagulation analyzer (Sysmex CA-500 Series Systems, Siemens, Breda, the Netherlands).

**SAFS**

Fibrinogen levels were measured using an automated clot-rate assay based upon the original method of Clauss[55] on the ST4 Instrument (Diagnostica Stago), with standardization with the College of American Pathologists (CAP) reference material.

**SardiNIA**

Fibrinogen levels were measured using the Clauss method[49].

**SHIP**

A non-fasting blood sample was drawn from the antecubital vein in the supine position and immediately analyzed or stored at -80°C. Plasma fibrinogen concentrations were assayed according to Clauss using an Electra 1600 analyzer (Instrumentation Laboratory, Barcelona, Spain)[49]. Coagulation time is measured and transferred into the result in g/L by applying a reference curve calculated in the laboratory. The assay proves linearity between 0.7 – 7 g/L. The analytical sensitivity of the assay was 0.7 g/L. Internal quality control measures were performed daily using two levels of manufacturers' control materials. External quality control measures were performed on a regular basis by participating in analysis programs. The inter-assay coefficients of variation were 4.61 % at low levels (mean value = 0.95 g/L) and 1.82% at high levels (mean value = 3.22 g/L) of control material.

**TwinsUK**

Fasting blood samples was taken from samples into 0.13 trisodium citrate containers (Becton Dickinson, Oxford, United Kingdom) at room temperature, centrifuged at 2560g for 20 minutes to obtain platelet-poor plasma within 1 hour of collection and stored at –40°C until analysis. Fibrinogen levels were determined using the Clauss method[49,56].

**VIKING**

Blood samples were drawn into sodium citrate tubes (Sarstedt) after an overnight fast. After centrifugation at 4°C, plasma was aliquoted and frozen at -40°C until analysis. Fibrinogen was measured using the Clauss assay[49].

**WGHS**

Fibrinogen in plasma from the baseline blood sample was measured by a mass-based immunoturbidimetric assay (DiaSorin) with reproducibility of 5.20% and 3.99% at concentrations of 0.99 and 2.74 g/L respectively.

**WHI**

Fibrinogen measurement in WHI was performed using a STA-R coagulation analyzer from Diagnostica Stago (16.6%), optical clot detection (80.1%), BNII nephelometer utilizing a particle enhanced immunoepholometric assay (1.4%), or an unknown method (1.9%).

#### 3. TOPMed Phenotype Harmonization

Measures of plasma fibrinogen for TOPMed participants with WGS data were harmonized to ensure that they were in the same units (g/L) and did not have unexpected distributions or an excess of outliers. Harmonization was performed for all participants of each study, not just those sequenced. After harmonization, duplicated samples (as identified by the TOPMed Data Coordinating Center (DCC)) were removed. These included technical replicates, within and between study duplicates, and monozygotic twins. Individuals without WGS data or with missing covariate information were also removed. Harmonized phenotypes were then uploaded to the Analysis Commons[57] for centralized genetic analysis.

#### 4. Whole-genome Sequencing of TOPMed Participants

TOPMed WGS methods have been described previously[58]. In brief, WGS was conducted at six sequencing centers at a mean depth of >30X using Illumina HiSeq X Ten instruments. Joint variant discovery and genotype calling were conducted by the TOPMed Informatics Research Center (IRC) across all TOPMed studies using the GotCloud pipeline, resulting in a single genotype call set encompassing all of TOPMed (TOPMed Freeze 6). Variant quality control performed by the TOPMed IRC consisted of the removal of variants failing the support vector machine filter, with excess heterozygosity or Mendelian inconsistencies, overlapping centromeric or other low complexity regions, or with missingness greater than 5%. Autosomal variants with mean sequencing depth < 10 were excluded. Sample quality control was performed by the TOPMed Data Coordinating Center (DCC) and consisted of the removal of duplicate samples, samples with sex discrepancies, misidentified samples, samples with consent issues, and samples with poor quality based on concordance of WGS and genotyping array data.

#### 5. Genotype Imputation of non-TOPMed Studies

Genotype array data for CHARGE studies and unsequenced TOPMed study participants were imputed to the densest available imputation panel. A total of 35 studies imputed to the TOPMed Freeze 6 reference panel[58] and four to the Haplotype Reference Consortium (HRC) reference panel[59]. Imputation was performed by each study individually using standard methods and following instructions provided centrally by the CHARGE Hemostasis Working Group based on the recommendations of the TOPMed DCC. HRC positions were mapped to hg19 and TOPMed Freeze 6 was hg38 therefore variant positions for studies with HRC imputation were converted to hg38 using liftOver[60] after genetic association analysis but prior to results quality control.

#### 6. Genome-wide Association Analyses and Meta-analyses

TOPMed WGS genetic analyses were conducted using inverse normalized and rescaled residuals adjusting for age, sex, population group*study, TOPMed sequencing phase, study-specific parameters, 11 ancestry informative principal components, and a kinship matrix. Single variant and aggregate gene-based tests were implemented using the SMMAT function of GENESIS with the Analysis Commons cloud computing platform[57,61]. Aggregate tests included only variants with minor allele frequency (MAF) <5% and minor allele count (MAC) ≥1 and used 3 strategies for variant selection: (1) loss of function (LOF), (2) LOF and deleterious missense (LDM), and (3) coding, enhancer, and promoter variants.

Studies without sequencing data undertook single variant analyses only within each population group using their software of preference and the same model described above. Summary statistics were provided for central meta-analysis.

Quality control of the single variant GWAS summary statistics was undertaken using the EasyQC package for R[62]. Variants were removed based on the following filtering criteria: estimated minor allele count (minor allele count x imputation quality; eMAC) < 6, absolute effect size (beta) > 5, standard error > 10, sample size < 30, or imputation quality < 0.30.

Meta-analysis was completed using GWAMA[63]. Genomic control was applied to each study individually but not to the meta-analysis results. Meta-analysis was completed (i) within each population group for just the CHARGE studies, (ii) just the TOPMed WGS studies, as well as (iii) the full TOPMed+CHARGE multi-population mega-analysis. 26,873,415 SNPs were included in the TOPMed+CHARGE multi-population mega-analysis and statistical significance was set at p<5.0E-09[64].

#### 7. Post-GWAS analysis

##### a. Conditional analysis

For all genome-wide significant regions from the TOPMED+CHARGE mega-analysis, conditional analyses were undertaken using “cojo-slct”[65] within GCTA[66]. All TOPMed WGS samples that contributed to the GWAS were used for the linkage disequilibrium (LD) reference panel. Options for cojo-slct were as follows: (i) minor allele frequency (MAF) cutoff = 0.0001, (ii) window size = 10 MB, (iii) collinearity cutoff=0.9, (iv) frequency difference threshold = 0.2, and (v) p-value threshold = 5.0E-09. Cojo-slct analysis was attempted using AFR-only and EUR-only ancestry-specific mega-analysis results and LD panels but power was severely reduced for AFR due to the small sample size, so results were not informative.

#### b. Variance Explained

Percent variance explained was estimated with summary level data reported by GCTA[66] for the 69 conditionally independent SNPs from the TOPMed+CHARGE mega-analysis, using the approximation derived by Shim et al[67] (see **Supplemental Materials S1 of Shim et al**).

### 8. Functional Annotation of Fibrinogen-Associated Variants

#### a. Variant Effect Prediction

Proxy variants based on linkage disequilibrium (LD) were obtained for the 69 conditionally independent SNPs from the TOPMed+CHARGE mega-analysis using TOP-LD[58,68] (http://topld.genetics.unc.edu/about.php) European and African ancestry reference panels (r^2>0.8). rsIDs for our SNPs and their LD-proxies in each region were uploaded to Ensembl Variant Effect Predictor (VEP)[69] (https://asia.ensembl.org/info/docs/tools/vep/index.html) to determine the nearest gene and the top Ensembl-predicted consequence for each variant. CADD PHRED[70,71] and LoFTool[72] predicted impact scores were obtained for all variants through VEP. For predicted coding variants, InterPro predicted position and amino acid substitution[73], SIFT[74], and PolyPhen[75,76] scores were obtained through VEP.

#### b. Overlap of Fibrinogen Signals to GWAS Catalog Associations

The NHGRI-EBI GWAS catalog v1.0.2, containing lead, significant (5.0E-08) variants from each uploaded GWAS study was downloaded October 29, 2022 (https://www.ebi.ac.uk/gwas/downloads). The same set of SNPs used for VEP annotation were queried in the GWAS catalog by rsID. We manually assessed all mapped traits to generate broad categories - we considered regions with a GWAS catalog signal for alkaline phosphatase, aspartate aminotransferase, alanine aminotransferase, gamma-glutamyl transferase, albumin, or sex hormone-binding globulin - as broadly overlapping with “liver enzymes”. Similarly, we considered regions with a GWAS catalog signal for platelet count, leukocyte count, neutrophil count, erythrocyte count, lymphocyte count, mean corpuscular hemoglobin concentration, mean corpuscular volume, myeloid white cell count, glycoprotein measurement, hematocrit, hemoglobin measurement, lymphocyte percentage of leukocytes, mean corpuscular hemoglobin, plateletcrit, red blood cell density measurement, neutrophil percentage of leukocytes, red blood cell distribution width, granulocyte count, HbA1c measurement, neutrophil count and basophil count, neutrophil count and eosinophil counts, neutrophil percentage of granulocytes, reticulocyte count, reticulocyte measurement, soluble triggering receptor expressed on myeloid cells 2 measurement, and/or total blood protein measurement as overlapping with “blood cell traits”.

#### c. Regulatory Annotation

All conditionally independent SNPs and their LD proxies were also queried by position (hg38) for overlap with Genehancer v5.12 regions, downloaded from Genecards on November 13, 2022[77] (https://www.genecards.org/).

To further assess which variants overlap with regions of open chromatin in liver tissue, we aligned variants to previously reported ATACseq (assay for transposase-accessible chromatin with high-throughput sequencing) consensus peaks determined using 20 healthy liver tissue samples by Currin et al[78]. ATACseq consensus peak regions (regions of DNA with an ATACseq peak in 3 or more samples) were downloaded from the Gene Expression Omnibus GSE164942. All SNPs and LD-proxies identified in our study were converted to hg19 coordinates using liftOver within the UCSC Genome Browser[60], then aligned with the ATACseq consensus peaks. We further compared our SNPs and LD-proxies to liver chromatin accessibility quantitative trait loci (caQTL) with effects in *cis*- (1 kb of target peak center) or *trans*- (within 100 kb of target peak center), as reported in Currin et al[78].

GTEx v8 conditionally independent expression QTL (eQTL) and splicing QTL (sQTL) datasets, mapped using stepwise regression, were obtained from the GTEx portal (https://gtexportal.org/home/datasets), and used to assess whether any fibrinogen lead variants or LD-proxy were predicted to alter expression or splicing of genes across all tissues mapped in GTEx.

#### 9. Transcription-wide Association Studies

We performed transcriptome-wide association study (TWAS) analysis using S-PrediXcan[79] to identify associations between *cis*-genetic components of gene expression and plasma levels of fibrinogen in mechanistically related tissues, namely artery (aorta, coronary, or tibial), liver, and whole blood. We used the prebuilt prediction models that were based on GTEx v8 multivariate adaptive shrinkage in R to estimate variants' weight on gene expression levels in chosen tissues[79–81]. Given that the reference models were created based on the European-ancestry population, we limited the analysis to GWAS results of European-ancestry individuals only. S-PrediXcan results of chosen tissues were then be leveraged using S-MultiXcan[82]. We determined significant TWAS signals using Bonferroni correction for the total number of genes in all models.

#### 10. Fine-mapping

We performed TWAS fine mapping using FOCUS in order to avoid false TWAS signals caused by co-regulation and the pleiotropic effects of SNPs at GWAS risk loci[83]. FOCUS models marginal TWAS z-scores of all genes in the same region considering SNP LD correlations and tagged pleiotropic effects of SNPs on the trait. Given generated z-scores, posterior inclusion probability (PIP) for a gene to be causal is derived and then used to form a credible set of putative causal genes. In this analysis, we used GTEx v8 MASH-R models as the source of eQTL weights and the European-based PROCARDIS database as the reference for LD correlations. PIP ≥ 0.95 was used as the threshold to determine putative causal genes in FOCUS results.

#### 11. Colocalization

We performed colocalization to identify shared variants between eQTL studies and GWAS to support gene-trait associations using fastENLOC. fastENLOC uses a Bayesian hierarchical model to generate SNP-level colocalization probabilities (SCP) and regional colocalization probabilities (RCP)[84,85] We took the precomputed GTEx multiple-tissue eQTL annotations and fibrinogen GWAS PIP input to perform fastENLOC for each of the 4 chosen tissues. We considered RCP > 0.5 as strong evidence of colocalization.

#### 12. VA Million Veteran Program (MVP) Phenome-wide Association Study (PheWAS) & Polygenic Risk Scores (PRS) analysis

Polygenic risk scores were derived using the independent variants identified by the GCTA analysis listed above. Three scores were derived, (i) weighting by the variant beta from the multi-population mega-analysis, (ii) weighting by the variant beta from the EUR-only meta-analysis, and (iii) weighting by the variant beta from the AFR-only meta-analysis. PRS were then standardized to standard deviation units. A phenome-wide association study (PheWAS) was performed for each of the three PRS within EUR and AFR participants of the VA Million Veteran Program[33] (MVP), for any ICD-based PheCode[86] that had more than 500 cases and controls. Logistic regression models were adjusted for age, sex, and the first five population-specific principal components. For sex-specific PheCodes, sex was removed from the model and values for the opposite sex were set to "NA". The number of independent PheCodes was calculated for each population (AFR=690; EUR=965) and Bonferroni threshold then applied to determine significance (AFR = 0.05/690 = 7.25E-05; EUR=0.05/965=5.18E-05). Additional sensitivity analyses were completed by creating the PRSs removing SNPs in the *FGG* gene region (rs2066874, rs148685782, rs28577061, rs149748987, rs6536024), or the fibrinogen gene cluster region (rs2227401, rs6054 + *FGG* SNPs).

#### 13. Determination of Independent PheCodes in VA Million Veteran Program

In order to determine the number of independent PheCodes tested, the phenotypic correlations were calculated for the PheCodes used in the MVP PheWAS. Principal components (PCs) were calculated from the correlations as well as the variance explained by each of the PCs. The number of independent PheCodes was determined by the number of PCs required to get a variance explained of 0.99.

### Supplemental Figures

#### Supplemental Figure 1 – Phenotype Harmonization in TOPMed

Untransformed

Residuals - Round 1

InvNorm, Rescaled Residuals

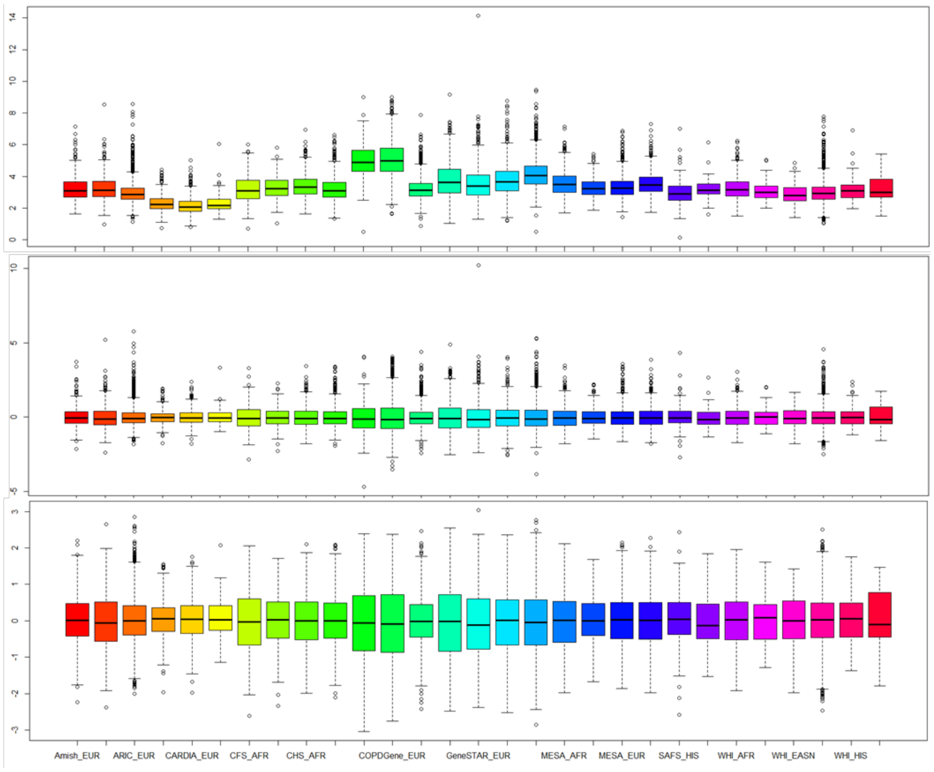

### Supplemental Figure 2 – Genetic Analysis Composition, Sample Size, and Workflow

#### Participating Studies

**TOPMed**
Freeze 6 Whole Genome Sequence (WGS)

| Population | N Studies | N Samples |
|---|---|---|
| AFR | 10 | 8,475 |
| ASN | 2 | 741 |
| EUR | 10 | 21,612 |
| HIS | 3 | 1,536 |
| **TOTAL** | **13** | **32,572** |

**CHARGE/TOPMed**
No WGS, TOPMed Imputed

| Population | N Studies | N Samples |
|---|---|---|
| AFR | 2 | 2,666 |
| ASN | 0 | 0 |
| EUR | 6 | 7,750 |
| HIS | 1 | 357 |
| **TOTAL** | **7** | **10,773** |

**CHARGE**
TOPMed Imputed

| Population | N Studies | N Samples |
|---|---|---|
| AFR | 1 | 142 |
| ASN | 0 | 0 |
| EUR | 27 | 115,562 |
| HIS | 1 | 168 |
| **TOTAL** | **28** | **115,562** |

**CHARGE**
HRC Imputed

| Population | N Studies | N Samples |
|---|---|---|
| AFR | 0 | 0 |
| ASN | 0 | 0 |
| EUR | 4 | 5,005 |
| HIS | 0 | 0 |
| **TOTAL** | **4** | **5,005** |

#### Final Analysis Sample Sizes

**Meta-Analysis**

| Population | N Studies | N Samples |
|---|---|---|
| AFR | 3 | 2,808 |
| EUR | 37 | 128,007 |
| HIS | 2 | 525 |
| **TOTAL** | **39** | **131,340** |

**Mega-Analysis**

| Population | N Samples |
|---|---|
| AFR | 11,283 |
| EUR | 149,619 |
| **Multi-Population** | **163,912** |

WGS = Whole Genome Sequencing; AFR = African or African American; ASN = Asian and Asian American; EUR = European or European American; HIS = Hispanic

#### Supplemental Figure 3 – Workflow for GCTA conditional analysis and MVP Phenome-wide Association Study (PheWAS)

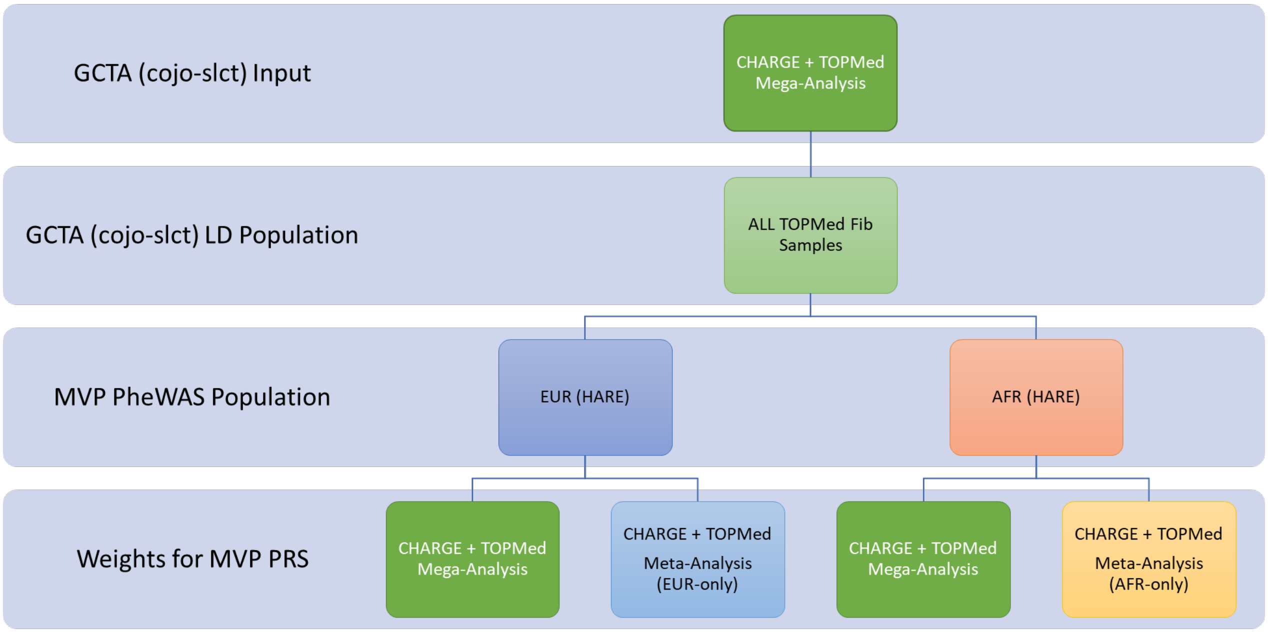

LD = linkage disequilibrium;  PheWAS = Phenome-wide Association Study; MVP = Million Veteran Program; HARE= Harmonized Ancestry and Race/Ethnicity; PRS = polygenic Risk Score; AFR = African American; EUR = European American

### Supplemental Figure 4 – MetaXcan Results from Various Tissues

#### A. All Tissues (MEGA)

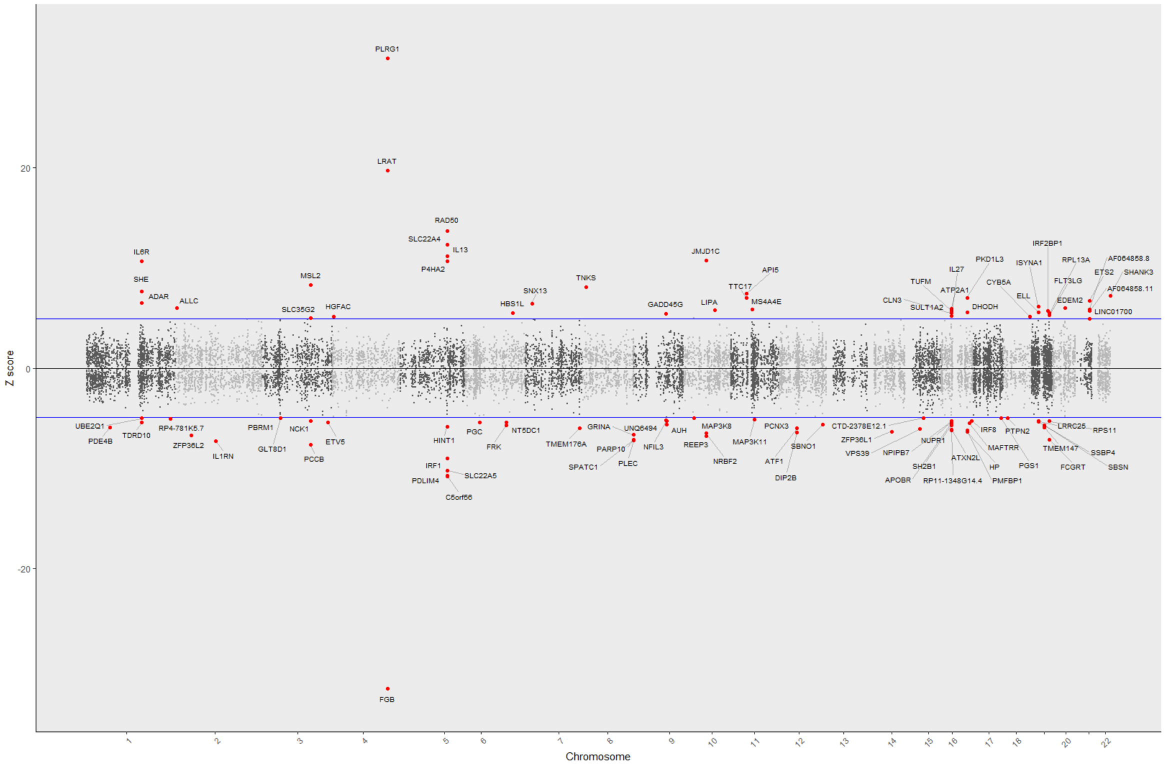

#### B. All Tissues (CHARGE-EUR)

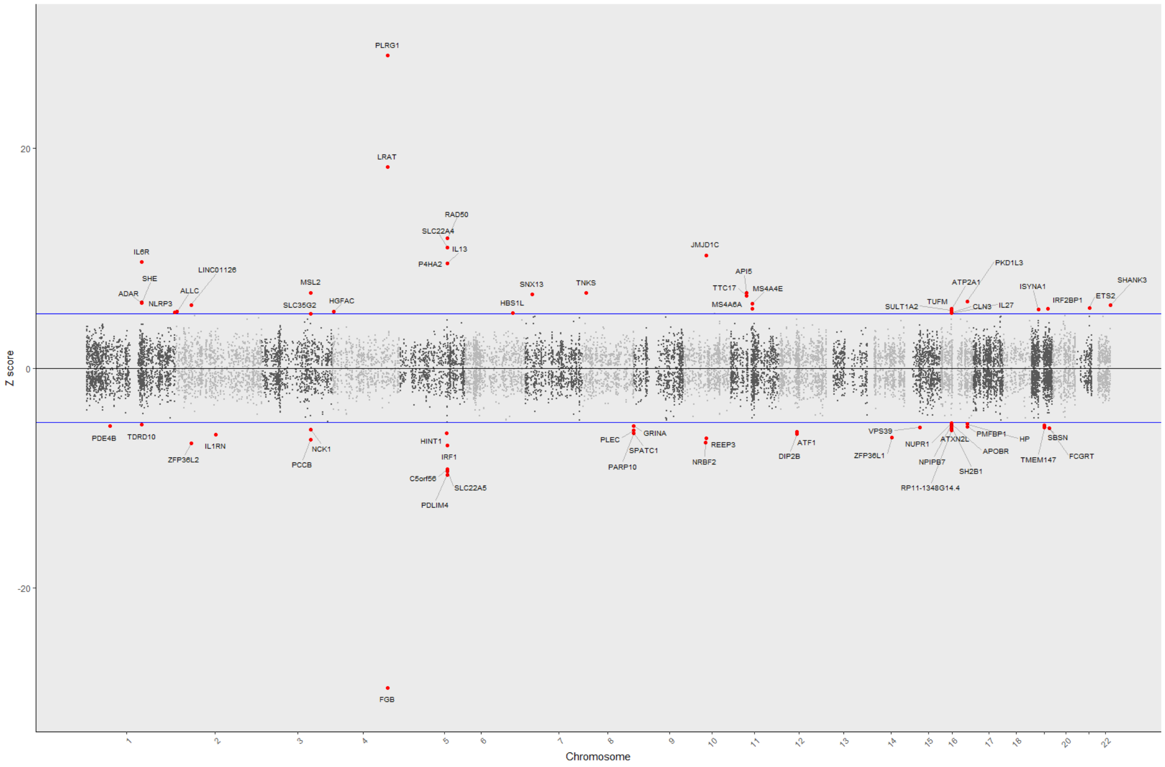

C. Artery - Aorta (MEGA)

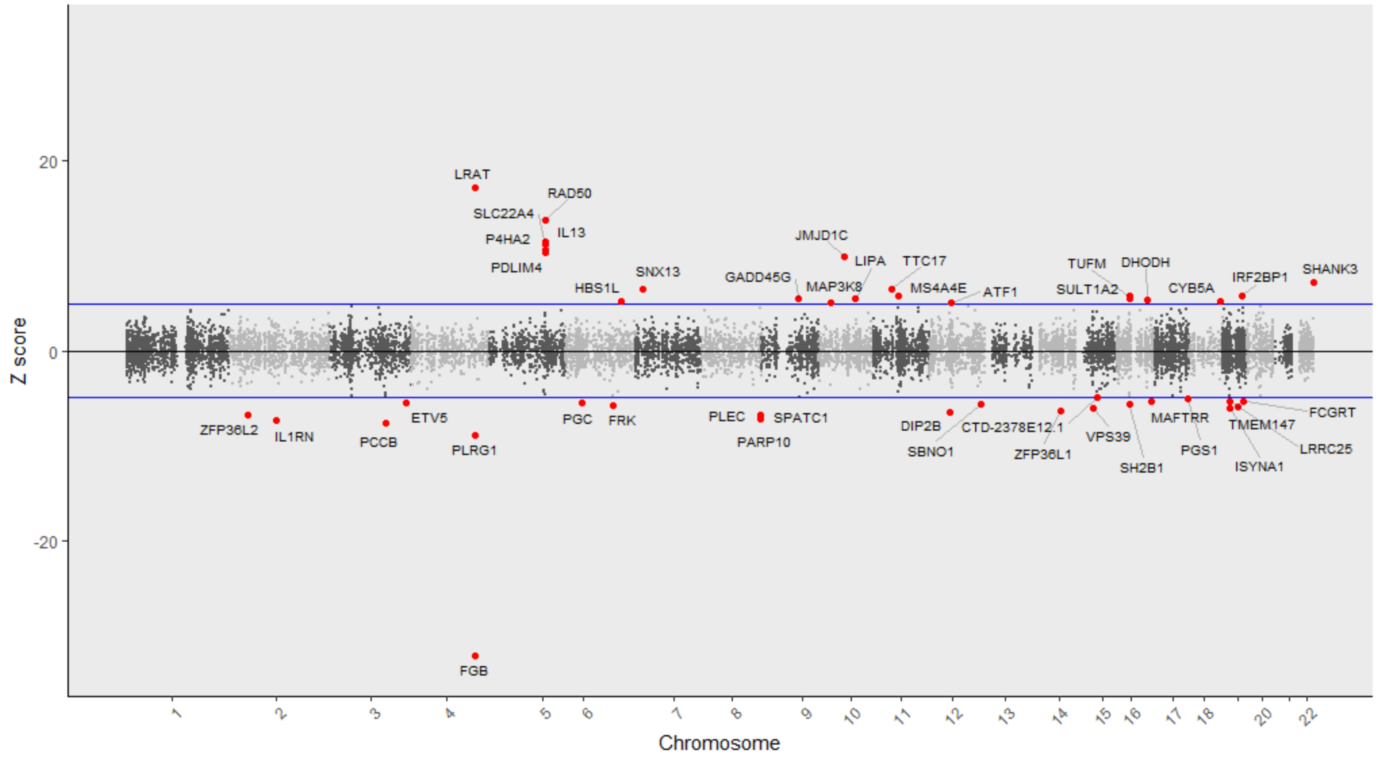

D. Artery - Aorta (CHARGE-EUR)

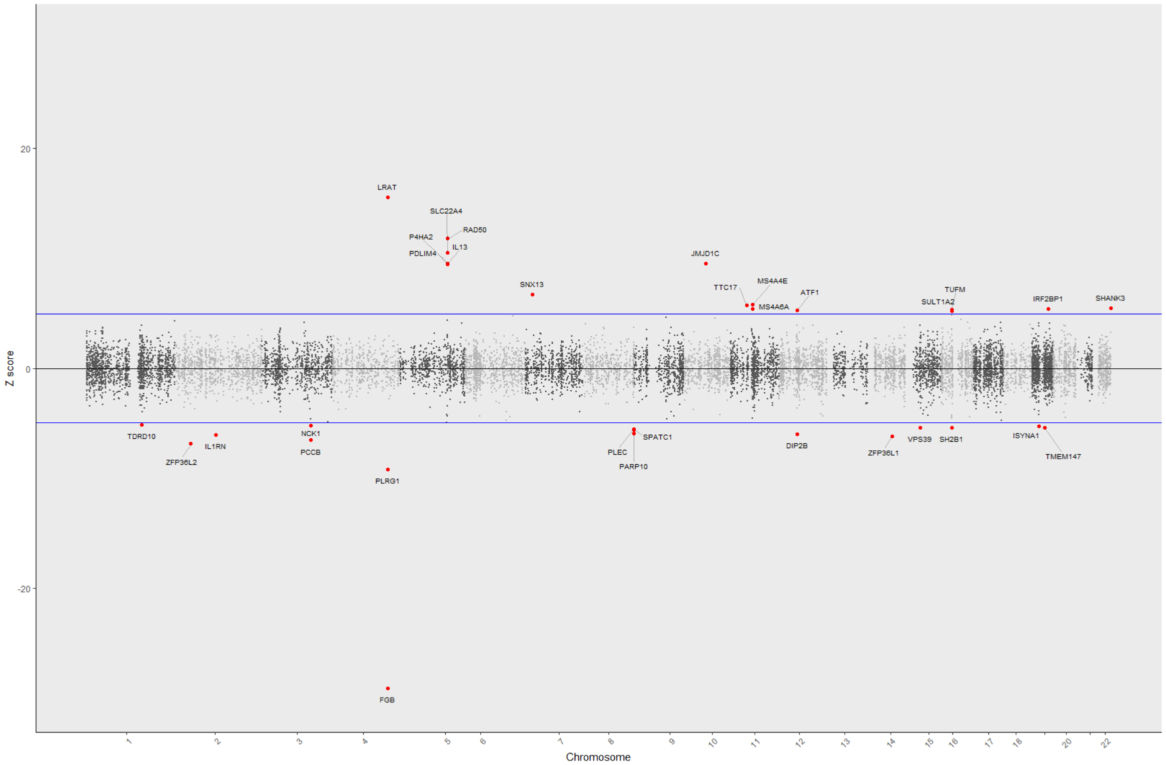

E. Artery - Coronary (MEGA)

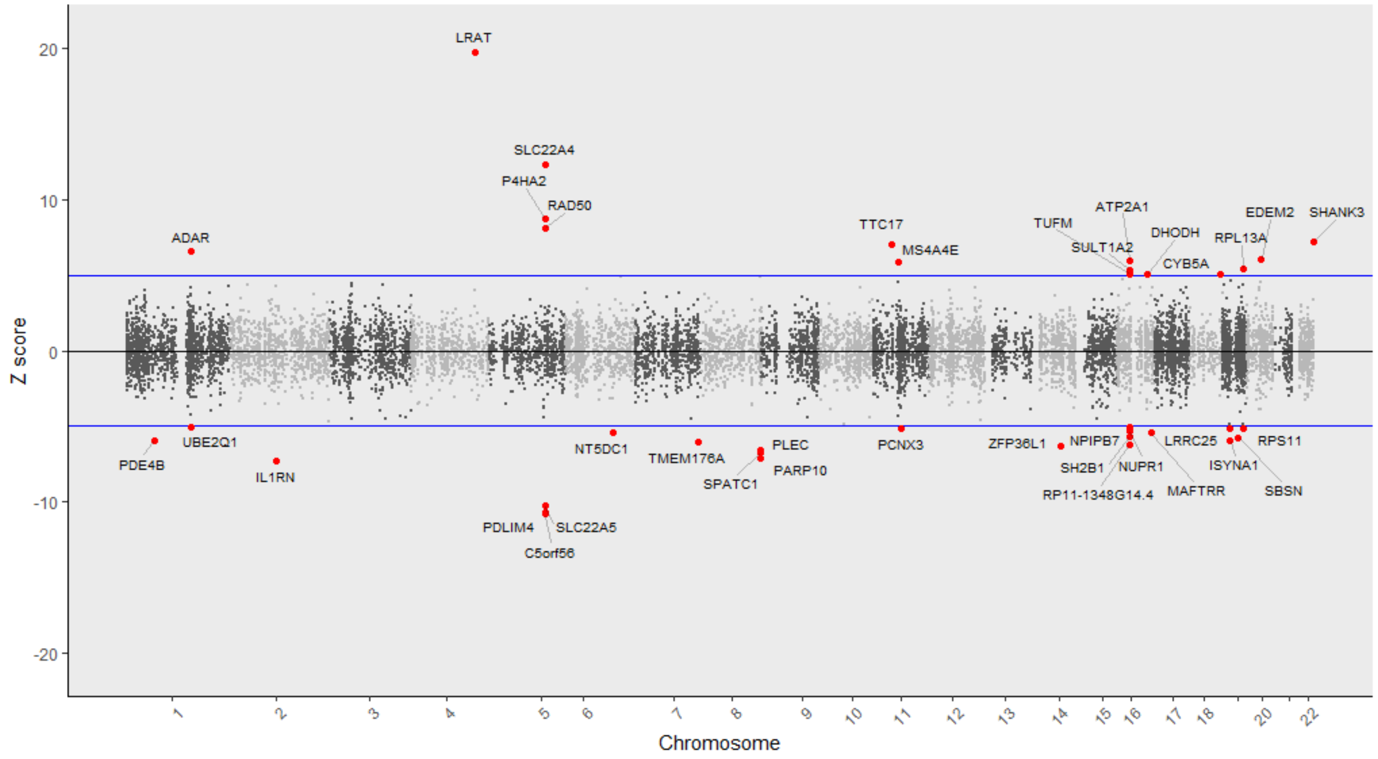

F. Artery - Coronary (CHARGE-EUR)

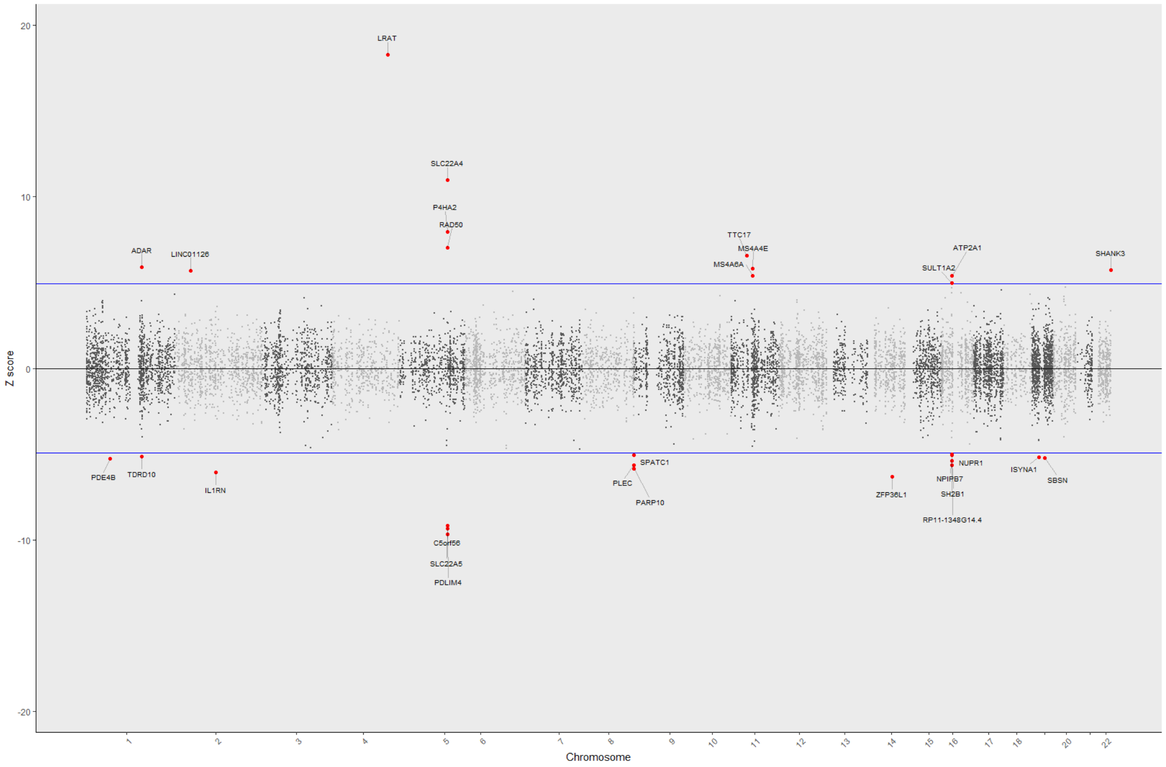

G. Artery - Tibial (MEGA)

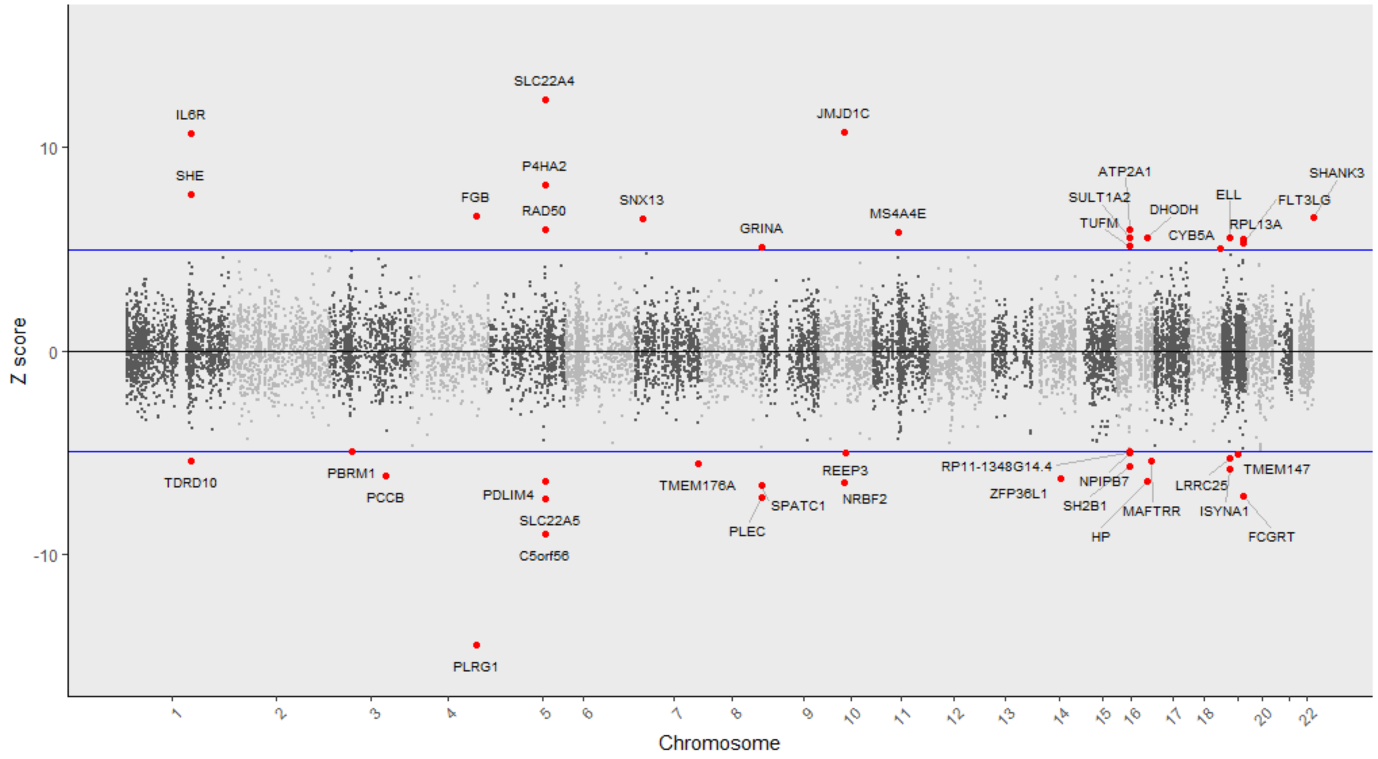

H. Artery - Tibial (CHARGE-EUR)

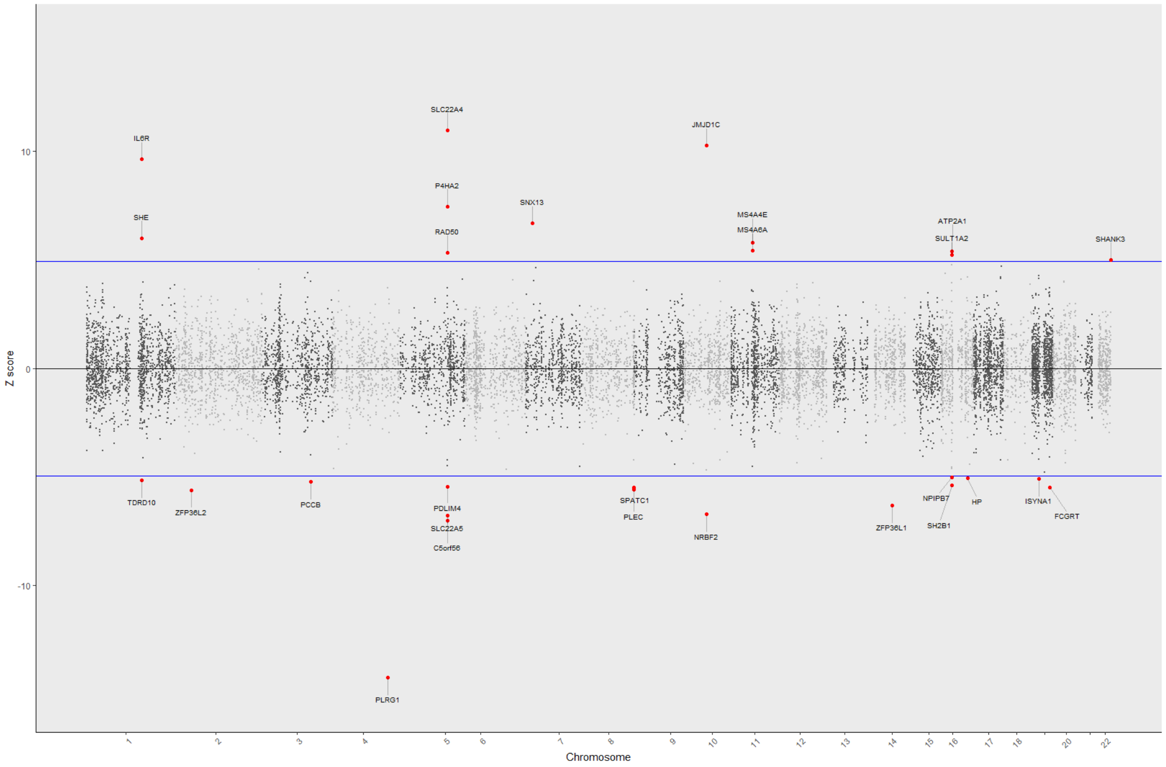

I. Liver (MEGA)

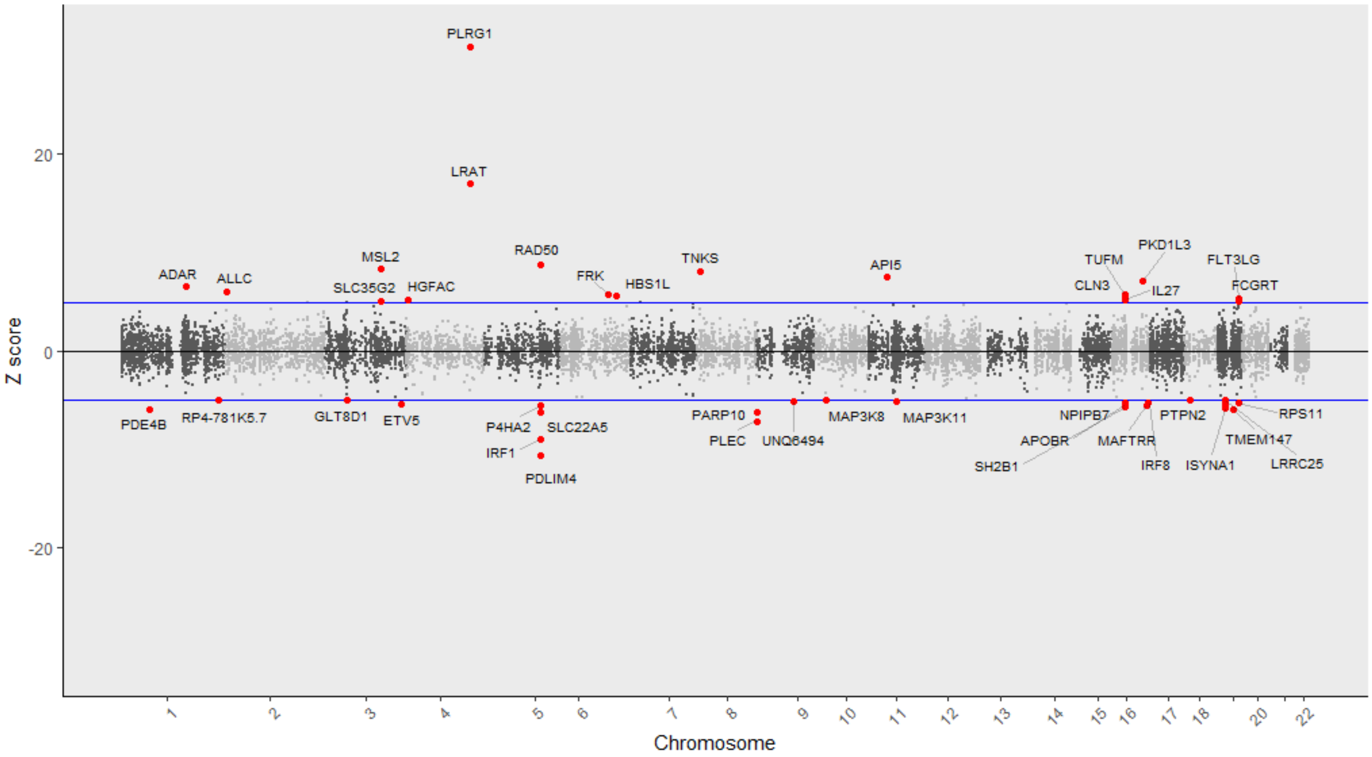

J. Liver (CHARGE-EUR)

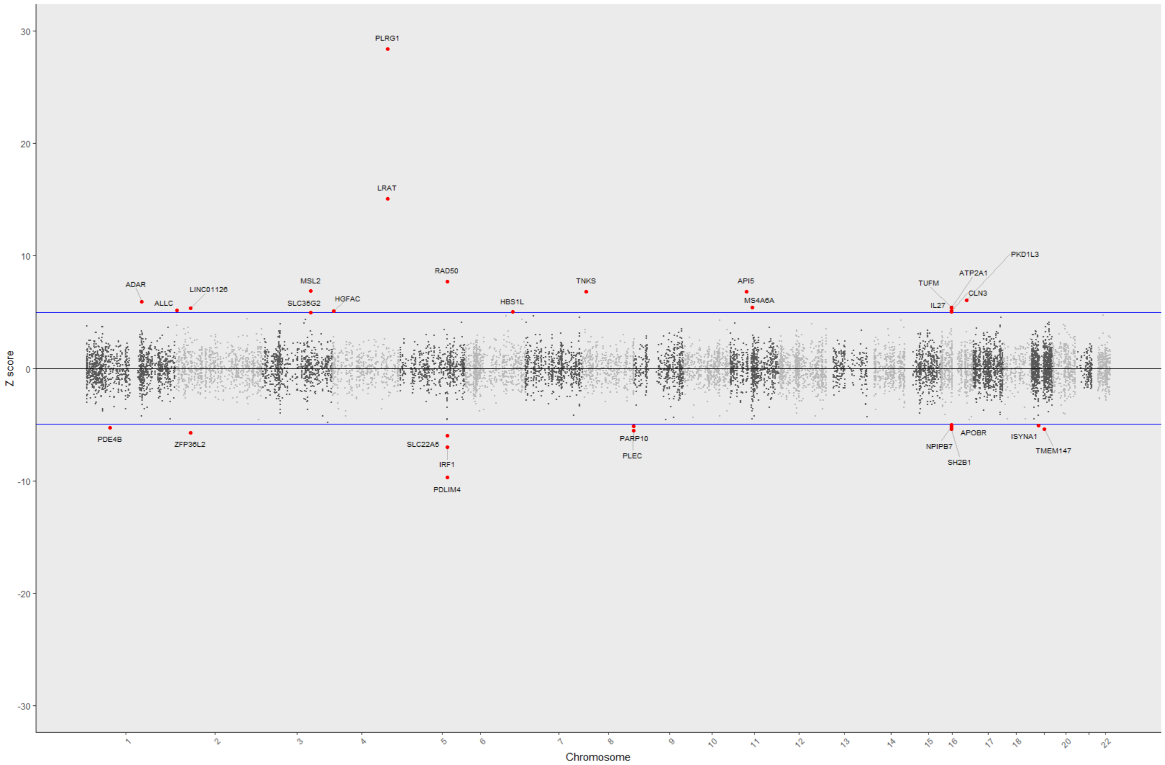

#### K. Whole Blood (MEGA)

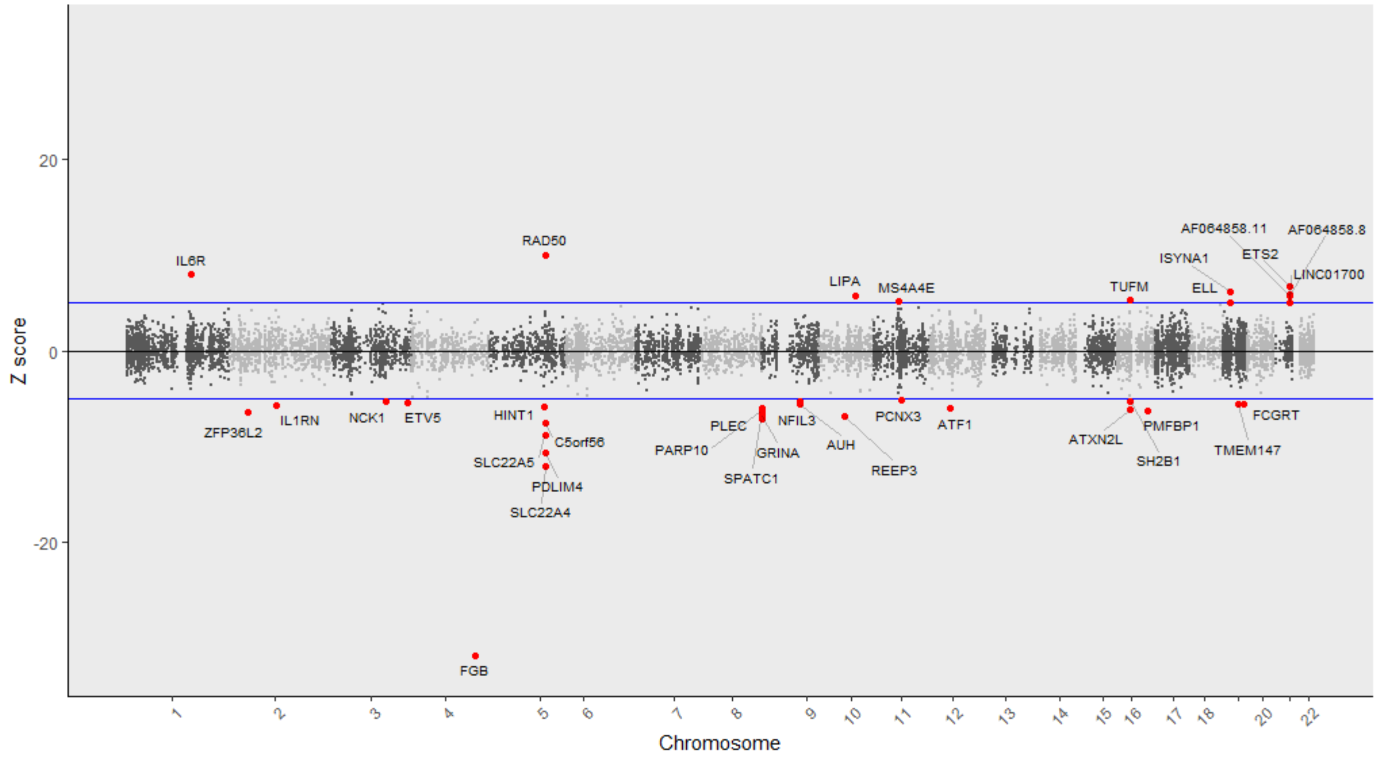

#### L. Whole Blood (CHARGE-EUR)

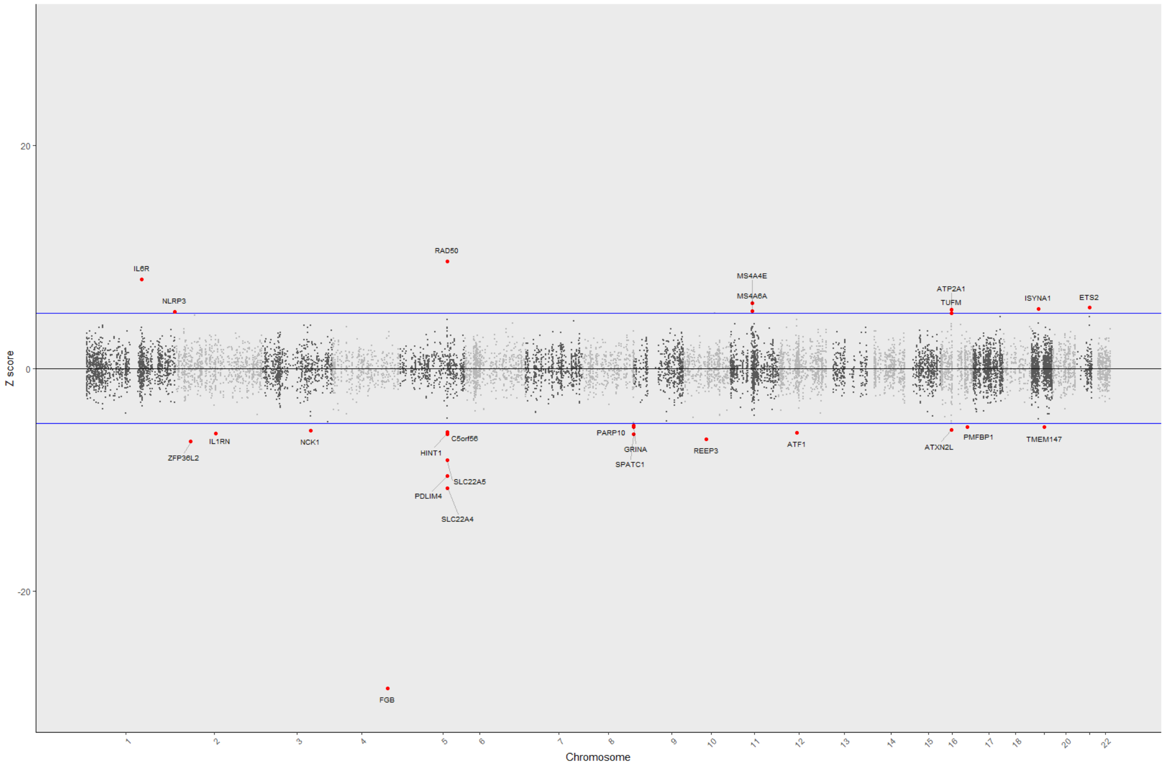

### List of Supplemental Tables

*See "TOPMed_CHARGE.Fibrinogen.Supplemental_Tables.June_1_2023.xlsx"*

**Supplemental Table 1** - Top SNP from TOPMed & CHARGE meta-analysis at each region reaching genome-wide significance (p<5E-09) for the single variant association with fibrinogen.

**Supplemental Table 2** - SNPs selected by GCTA (cojo-slct) using TOPMed multi-ancestry reference panel for LD.

**Supplemental Table 3** - LD between top SNPs and associations from previous publications.

**Supplemental Table 4 -** Genes significantly associated with fibrinogen levels in TOPMed WGS aggregation tests.

**Supplemental Table 5** - Summary of Annotations per Signal.

**Supplemental Table 6** - Full Results of Variant Annotations for Conditionally Distinct SNPs and LD Proxies.

**Supplemental Table 7** – SNP-eQTL variant colocalization

**Supplemental Table 8** – SNP-eQTL regional colocalization

**Supplemental Table 9** – Prioritized genes in transcriptome-wide association fine-mapping analysis implemented in FOCUS.

**Supplemental Table 10** - Prioritized genes in transcriptome-wide association fine-mapping analysis implemented in FOCUS.

**Supplemental Table 11** - SNPs and Weights for Polygenic Risk Score Analysis in MVP.

**Supplemental Table 12** - Associations of Polygenic Risk Score SNPs with Statistically Significant PheCodes in MVP.

**Supplemental Table 13** - PRS Sensitivity Analysis for significant PheCodes removing (i) SNPs in FGG region and (ii) whole of fibrinogen gene cluster region.
